# Synthetic cannabinoids consumed via e-cigarettes in English schools

**DOI:** 10.1101/2024.08.12.24311617

**Authors:** Gyles E. Cozier, Matthew Gardner, Sam Craft, Martine Skumlien, Jack Spicer, Rachael Andrews, Alexander Power, Tom Haines, Richard Bowman, Amy E. Manley, Peter Sunderland, Oliver B. Sutcliffe, Stephen M. Husbands, Lindsey Hines, Gillian Taylor, Tom P. Freeman, Jennifer Scott, Christopher R. Pudney

**Affiliations:** Department of Life Sciences, University of Bath, Bath, BA2 7AY, UK; Department of Psychology, University of Bath, Bath, BA2 7AY, UK; Department of Social and Policy Science, University of Bath, Bath, BA2 7AY, UK; Department of Computer Science, University of Bath, Bath, BA2 7AY, UK; School of Physics and Astronomy, University of Glasgow, Glasgow, G12 8QQ, UK; Bristol Medical School, University of Bristol, Bristol, BS8 2PS, UK; MANchester DRug Analysis & Knowledge Exchange (MANDRAKE), Department of Natural Sciences, Manchester Metropolitan University, Manchester, M1 5GD; School of Health and Life Sciences, Teesside University, Middlesbrough, TS1 3BX; Centre for Academic Primary care, Bristol Medical School, University of Bristol, Bristol, BS8 2PS, UK; Centre for Therapeutic Innovation, University of Bath, Bath, BA2 7AY, UK

**Keywords:** Synthetic cannabinoids, spice, K2, school, e-cigarette, THC

## Abstract

Synthetic cannabinoids (SCs), colloquially spice or K2, are the most common drug to be found in prisons in the UK, where they are associated with nearly half of non-natural deaths. In the community, SCs are associated with poly-drug users who are also likely to be homeless. People who use SCs report debilitating side effects and withdrawal symptoms, coupled with dependence. Until now, SC use was believed to be largely restricted to prison and homeless populations. However, media reporting in the UK has increasingly identified cases of children collapsing in schools, which are claimed to be associated with vaping and putatively the vaping of a drug, variously reported as tetrahydrocannabinol (THC) ‘synthetic cannabis’ or ‘spice’. We therefore conducted the first study to identify and quantity SCs in e-cigarettes routinely collected from schools. We sampled 27 schools from geographically distinct regions of England, representing a very broad range of social metrics (free school meals, persistent absenteeism, and SEN). The material was sampled by self-submission by individual schools of e-cigarettes seized during normal school operation and transferred to us for analysis via local police forces. We found a remarkably consistent picture where SCs were detected in 17.5 % of all e-cigarettes sampled, and in 21 of 27 (78 %) of all sampled schools. Moreover, the percentage of SC e-cigarettes positively correlated with a metric of social deprivation, the fraction of pupils eligible for free school meals. The SC positive e-cigarettes were almost entirely found in e-cigarette liquid bottles and refillable e-cigarette devices, with very few identified in single use e-cigarette products. Within the positive samples we found an average SC concentration of 1.03 mg mL^-1^ with a maximum of 3.6 mg mL^-1^. In contrast to the high prevalence of SCs, few samples contained THC (1.6 %). We suggest that pupils are being sold SC e-cigarettes as ‘cannabis’ and may be unaware they are consuming (and sometimes supplying) considerably more harmful drugs. Our findings are immediately crucial to policy policing and healthcare in the UK as well as to educational bodies and schools.

---

Synthetic cannabinoids (SCs), often referred to as spice or K2, are a class of synthetic drugs. At the time of writing there are over 300 known SCs and the prevalence of different SCs can change rapidly.^1,2^ SCs are considered to generally be highly potent and with high efficacy, often acting as full cannabinoid receptor agonists. However, their structure is dissimilar to typical cannabinoids found in cannabis, such as tetrahydrocannabinol (THC), which acts as a partial cannabinoid receptor agonist and has a considerably lower risk profile compared to SCs. Potential consequences of SC use include psychosis, seizures, hypertensive crisis, and death. There is only sparse research into the correlation between different SC structures and their pharmacology and risk profile. Indeed, there are now a number of studies that point to SCs potentially having effects at sites other than cannabinoid receptors.^3-5^

In the UK, data suggest that use of SCs in the general population is extremely low. For example, in the Crime Survey for England and Wales, SCs are subsumed under the broader category of Novel Psychoactive Substances (NPS), and in 2023 only 0.4% of adults (aged 16-59) reported use within the past-year.^6^ However, SCs are the dominant drug used in the British prison system^7,8^ and are commonly used by people who are homeless.^9^ That is, these drugs are typically associated with at-risk individuals and people with complex poly-substance use histories. People who use SCs report highly variable and unpredictable effects, which increases the risk of ‘going over’ – collapsing and becoming comatose.^10^ Indeed, nearly half of all non-natural deaths in British prisons have been associated with SC use.^10^ Additionally, many report strong withdrawal symptoms upon discontinuing use, which are common and more severe than cannabis withdrawal symptoms.^9, 11,12^

The mode of SC use varies significantly across different groups. In prisons, SC-soaked paper is inserted into a prison-issued e-cigarette device and the paper combusted, essentially smoked.^13,14^ In homeless communities, SC use is commonly via smoking herb-soaked material, somewhat resembling cannabis.^9^ There is little understanding how the mode of use affects SC toxicity, however in both of the modalities above, there is a high risk of ‘hot-spots’; locally high concentrations of SCs that might contribute to the risk profile of SC use, outside of their generally high potency.

The use of e-cigarettes has become common in England, with one in five 15-year-olds and 9% of 11-14 year olds reporting using e-cigarettes (‘vapes’).^15^ Coincident with the increasing use of e-cigarettes, there have been growing reports of SCs being found in e-cigarette liquid. Between January 2023 and April 2024, testing by the UK drug checking service, WEDINOS, showed that 41% of 122 submitted e-cigarettes contained SCs.^16^ Crucially, none of these samples were submitted with the purchase intent of SCs.

SC e-cigarette liquid is inexpensive, being sold for as little as £1.6 mL^-1^ and easily available online.^16^ Recent studies suggest the concentration of the SC e-cigarettes varies from ∼1 mg mL^-1^ up to a maximum reported value of 24.1 mg mL^-1^.^18^ Though we acknowledge that there are relatively few studies addressing the concentration range that is present. In the last year, there have been over 16 media reports of putative SC/THC related adverse effects in British schools (Figure 1) and we are aware of many more not reported by the media (personal communication from police forces across the UK). There is a fundamental lack of analytically confirmed data, and at scale, to assess the occurrence of SC e-cigarettes in schools.

**Figure 1.**
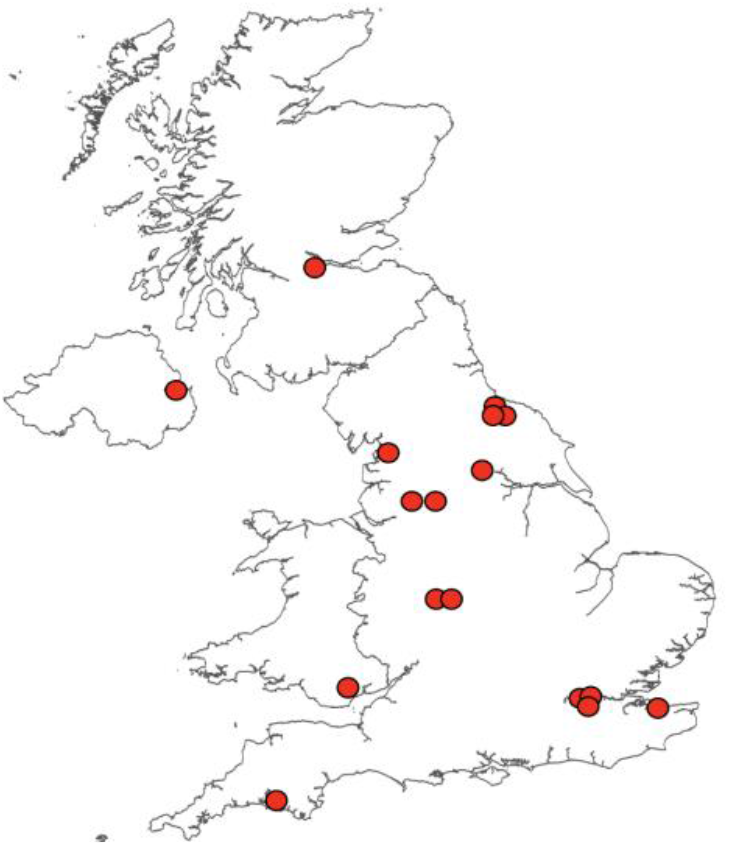
Media reports of adverse effects in schools associated with vaping and putatively associated with SC use.

THC overdose requires consumption of very high quantities, suggesting that the reported adverse effects are caused by something else. The risk of emergency medical treatment is estimated to be 30-fold greater for use of SCs compared to THC.^19^ Therefore, we hypothesise that SC e-cigarette may be commonly found in e-cigarettes used by school children. Herein, we sample seized e-cigarettes from secondary (age 11-18) schools across a range of regions in England, assaying for the presence of substances controlled under the Misuse of Drugs Act (1971) and the Psychoactive Substances Act (2016) and their concentration. Further, we demonstrate how a recent innovation in portable SC detection can be effectively used to monitor the presence of SC e-cigarettes at a local level.

## Results

To assess the presence and dosage of e-cigarettes containing SCs, we have sampled a number of different regions in England, designated R1 – R3. We have anonymised the specific regions sampled at the request of the police forces involved in the sampling. The sampling procedure is described in Materials and Methods. Table 1 shows key characteristics of the schools in the regions from UK Government reported data.^20^ From Table 1, the number of schools sampled, the fraction of pupils eligible for free school meals (FSM), persistent absence, special educational needs (SEN) provision and students with an education, health and care plan (EHCP) are all similar. That is, these regions are meaningfully comparable, and the combination of their statistical data is warranted. Similarly, the school metrics track well with the English national averages. We note that the fraction of FSM and persistent absence is slightly higher than the English mean at the time of writing – 30.5 ± 0.6 % *versus* 27.1 % and 28.8 ± 0.7 % *versus* 26.5 %, respectively.^20^ The difference is not over large given the absolute magnitude of the values, and combined with the other metrics suggest a reasonable comparison to the national picture. The individual data for schools is given in Tables S1-4. We have not used a postcode-based metric of deprivation, as schools tend to draw widely across an area and so we did not feel this would be useful or representative.

**Table 1.** The prevalence of occurrence of SC and THC in schools from selected regions.

| | R1 | R2 | R3 - In-field testing<sup>c</sup> | Combined average ± stdev |
| --- | --- | --- | --- | --- |
| No. of schools | 13 | 9 | 5 | 27 total |
| FSM; Ave. / England<sup>a, b</sup> | 30.6 / 27.1 % | 31.1 / 27.1 % | 29.9 / 27.1 % | 30.5 ± 0.6 % |
| Persistent absence; Ave. / England<sup>a, b</sup> | 28.2 / 26.5 % | 28.6 / 26.5 % | 29.5 / 26.5 % | 28.8 ± 0.7 % |
| SEN support; Ave. / England<sup>a, b</sup> | 12.0 / 12.4 % | 12.8 / 12.4 % | 13.1 / 12.4 % | 12.6 ± 0.6 % |
| EHCP; Ave. / England<sup>a, b</sup> | 2.3 / 2.4 % | 2.0 / 2.4 % | 2.5 / 2.4 % | 2.3 ± 0.3 % |
| Total samples | 156 | 270 | 84 | 510 |
| Total tested | 119 | 251 | 84 | 454 |
| Total SC - % total | 35 – 22.4 % | 40 – 14.8 % | 14 – 16.7 % | 89 – 17.5 % |
| Total THC | 1 – 0.6 % | 0 – 0 % | 7 – 8.2 % | 8 – 1.6 % |
| SC |  |  |  |  |
| Single use - % (total) | 1 (106) | 7 (196) | 0 (37) | 8 – 1.6 % |
| Liquid/refill - % (total) | 34 (50) | 33 (74) | 14 (47) | 81 – 15.9 % |
| Labelled liquid | 3 | 0 | 0 | 3 – 0.6 % |
| Unlabelled liquid | 11 | 16 | 9 | 36 – 7.0 % |
| Refillable device | 20 | 17 | 5 | 41 – 8.2 % |
| THC |  |  |  |  |
| Single use - % | 1 | 0 | 3 | 4 – 0.8 % |
| Liquid/refill - % | 0 | 0 | 3 | 3 – 0.6 % |
| Labelled liquid | 0 | 0 | 0 | 0 |
| Unlabelled liquid | 0 | 0 | 0 | 0 |
| Refillable device |  |  | 3 | 3 – 0.6 % |

a, Data obtained from ref 20. b, England state funded schools. c, data collected as described in the text.

In all cases the sampling was via a police coordinated submission of seized e-cigarettes, and brought to our laboratory for testing or in the case of R3 (described below), at a local police station. We stress that the submissions were unbiased, in that no specific schools were targeted, no specific selection protocol for identifying samples was defined and the seized material was identified by working teachers in each school, based on their usual seizure of e-cigarettes and liquids. Participation by schools was voluntary and at the request of the local police force. We have explicitly excluded samples provided by the police that already have intelligence associated that they are an SC e-cigarette or liquid. The seizures ranged in collection date from September 2023 – June 2024, effectively a school year. In total, 510 samples were collected (R1 = 156, R2 = 270, R3 = 84). That is, the data reported in Table 1, represents as close as is possible, an unbiased sampling of e-cigarettes.

First, we describe the data from R1 and R2 where the samples were returned to our laboratories and analysed by LC-MS and NMR. From R1, e-cigarette liquid could tractably be extracted from 119 / 156 submitted samples. From R2, liquid could tractably be extracted from 251 / 270 submitted samples. Each of these samples was then analysed by LC-MS, and for those positive for a substance controlled under the Misuse of Drugs Act (1971) and the Psychoactive Substances Act (2016), the concentration was then determined by NMR as described in Materials and Methods.

Figures S1 and S2 show representative LC-MS data for the positive samples, with the parent molecules and fragments ions used to identify compounds listed in Table S5. The corresponding structures shown in Figure S3. Figures S4-S7 show example NMR spectra used for quantitative analysis and the corresponding standard concentration plot shown in Figure S9.

Table 1 gives a summary of the resulting frequency of controlled drug identification and across different categories of sample, including single use (disposable) e-cigarettes and refillable device / liquids. Tables S1-S4 shows the data for all samples broken down to the individual school level. Figure 2 shows an image for all SC positives and Figure S9 shows an image for all THC positives. We define a ‘single use’ e-cigarette as one where it is not a design feature of the e-cigarette to be refilled. We note that some e-cigarettes, whilst not designed to be refilled do have the ability to be recharged, as the volume of liquid that can potentially be consumed exceeds the batteries capacity from a single charge.

**Figure 2.**
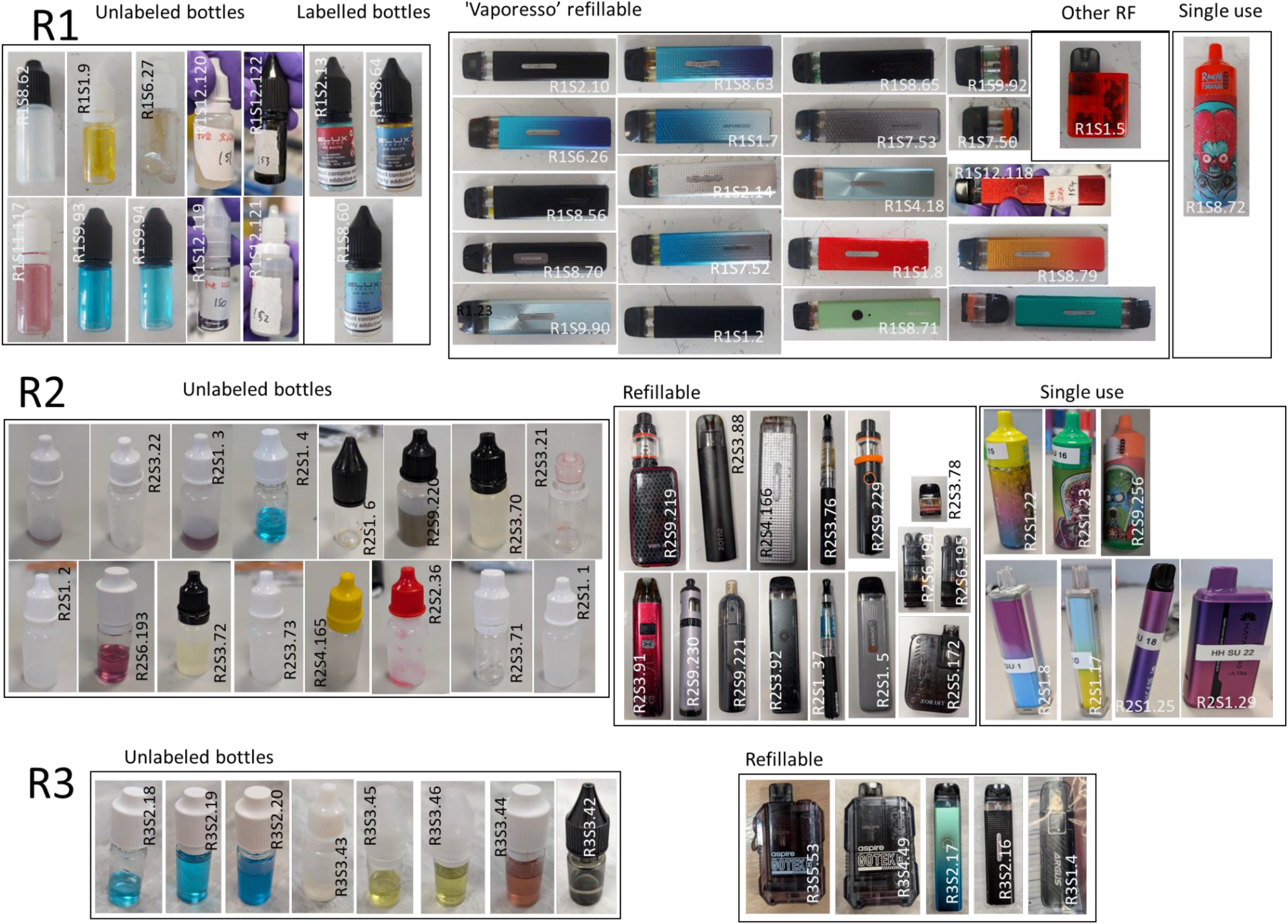
Presentation of SC e-cigarettes from each sampling exercise.

For R1, SCs were identified in 22.4 % of all e-cigarettes. The data show that only 1 single use e-cigarette contained SCs (0.6 %) and 1 contained THC (0.6 %). For R2, SCs were identified in 14.8 % of all e-cigarettes. SCs were found in 7 (2.6 %) single use e-cigarettes, though these were largely from a single school (R2.S1; 6 of 8 recorded positives in the study). No THC was identified. These data suggest that SCs are present primarily in liquid/refillable device and more rarely in single use e-cigarettes. We discuss THC in more detail below owing to its legal status in some countries and associated commercial product availability. We note that whilst it is possible to open single use e-cigarettes and replace / add to the liquid inside, it can be technically laborious and so the presence in liquids and refillable devices is entirely logical.

Having identified a high prevalence of SCs using LC-MS and NMR (which could not feasibly be implemented rapidly at scale), we wished to further demonstrate that SC e-cigarettes are in fact common in schools by sampling discrete schools in various regions of England, not covered by R1 and R2 above. We have recently reported a portable device for the generic detection of SCs^21^ and we have expanded this to the detection of THC from sealed e-cigarettes and e-cigarette liquids.^22^ This technology self-actuates an e-cigarette and extracts a small amount of vapour, condensed on a solid matrix with subsequent detection based on combined fluorescence and photochemical discrimination. We reasoned this tool would be useful to expand data sampling.

The device has the potential for the accuracy to be tuned to increase/decrease sensitivity with the limit of detection (LOD). As such, using the sample sets from R1 and R2 for calibration, we have tuned the device to have an LOD of 0.3 mg mL^-1^, with a corresponding accuracy of 95 %, specifically with e-cigarette liquids (usage statistics given in Table 2). Clearly, the device will miss a fraction of positive samples (below the set LOD). That is, whilst an imperfect tool, the device will only underestimate the occurrence of SCs and so we feel that balance is reasonable.

**Table 2.** Usage statistics of the presumptive device.

| FP | FN | TP | TN | Tot. |
| --- | --- | --- | --- | --- |
| 2 | 3 | 48 | 42 | 95 |
| Sensitivity (%) |  | Specificity (%) |  | Accuracy (%) |
| 94 |  | 95 |  | 95 |

With this tool in hand, we have sampled 5 additional schools using the portable device and the corresponding data are given in Table 1 under ‘R3 - in-field testing’. Similar to R1 and R2 these schools represent submissions from schools to the police and not with samples that are considered suspicious. These schools are drawn from two separate metropolitan regions (one and four schools), distinct from R1 and R2, that are 90 miles apart. The testing in each case took place in a police station, chaperoned by a police officer. The school metrics for these samples, given in Table 1, are similar to R1 and R2 and again suggest the comparison to the data set at large is meaningful. Indeed, these data show a similar percentage of SC presence as the other regions, being 16.7 % with the positive samples being shown in Figure 2. No positives were identified from single use e-cigarettes, potentially because of the LOD of the device and so we caution that the real positive rate may be higher than reported for R3.

R3 differs from R1 and R2 in that 6 THC e-cigarettes were identified (only 1 from R1 and R2 combined), shown in Figure S1. In 4/6 cases, the THC e-cigarette was a commercial product, apparently originating in the USA with a cost of ∼£15-65. In 2/6 cases the e-cigarette was designed to be filled with THC oil or resin and are marketed for this purpose on available web shops. The THC resin is clearly identifiable by eye and odour, versus the entirely clandestine SC e-cigarettes found in all other positive samples.

Combining the data from R1, R2 and R3, we find that the average percentage of all e-cigarettes containing SCs is 17.5 % (89 / 510). Indeed, the percentage of SC e-cigarettes is similar between R1-R3 being, 22.4, 14.8 and 16.7 %, respectively. That is, the percentage of e-cigarettes appears entirely consistent even in geographically distinct regions of England. This percentage is nuanced by the presence of very low concentrations and we discuss this below. From Table S1-3, the majority of positive samples contain MDMB-4en-PINACA, being present in 95% of SC positive samples. Other SCs identified include MDMB-PINACA, ADB-BUTINACA, MDMB-BUTINACA, MDMB-INACA and 4F-MDMB-BINACA. Of particular note, we found that sample R2S2.36 contained an SC but also a low concentration (0.1 mg mL^-1^) of heroin. Figure 2 shows an image of every SC positive sample identified.

Figures 3A and 3B show the combined profile of SC containing samples, including form factor and liquid colour. From Figure 3A, over half of the samples present as ‘normal’ e-cigarette liquid colouring (clear, yellow, brown), with the remaining being ‘uncommon’ colours for branded commercially available e-cigarettes (eg green, blue, pink). From Figure 3B, the vast majority of the positive samples are from bottles of liquid or from refillable devices, with 40.4 % of those samples being unlabelled bottles of liquid and 46.1 % being from refillable devices. SCs are also found in labelled (commercial packaging) bottles of liquid, but this is relatively rare, with only three samples being found in R1(Figure 2; R1S8.60, 64 and R1S2.13). We do not suggest these commercial products contained SCs at source, and they could trivially have SCs added, or be filled with an alternative liquid after purchase. Similarly, the presence of SCs in single use e-cigarettes is low, 1.6 % of all samples. It is interesting to note that 7 of the 8 SC positive single use e-cigarettes contain an illegally (in the UK) large amount of e-liquid (> 2 mL). Finally, from Figure 2, there is a regional tendency for the refillable devices to be of a specific commercial brand (Figure 2; R1.11-R1.29), but this is not found in R2-3, where there is a diversity of brands. However, there is a tendency for the refillable devices to have removeable caps that contain the liquid.

**Figure 3.**
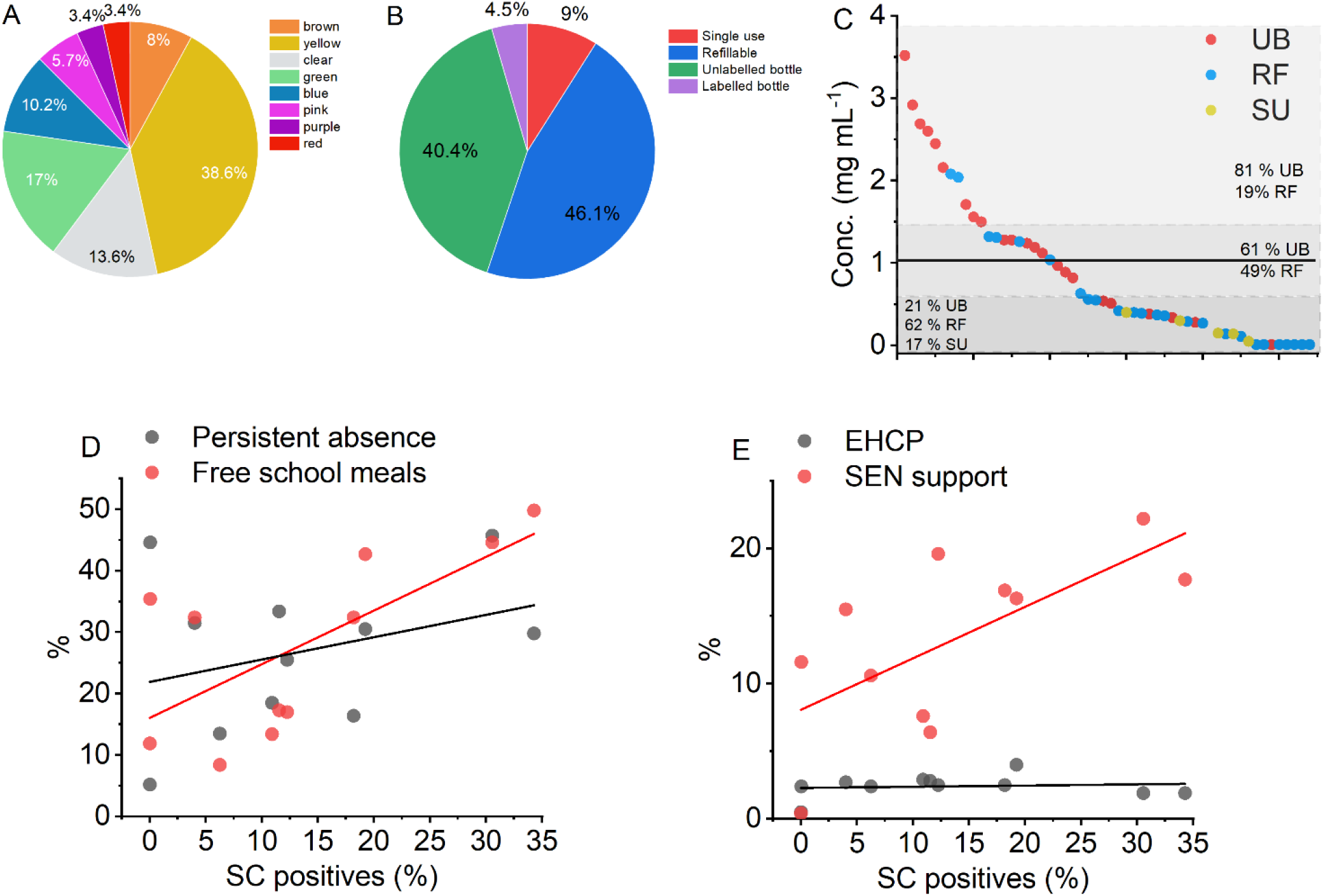
**A-C**, Physical presentation, and concentration distribution of SC e-cigarettes. The black solid line in panel C represents the average recorded concentration. The grey boxes highlight specific regions referenced in the text. The magenta dashed line shows the LOD for the presumptive device and the black dashed line shows the LOD for our NMR quantification methodology. **D-E**, Correlation with school metrics. Solid lines show the fit to a simple linear function and statistical correlations are discussed in the text.

We have determined the concentration of SCs present in the samples by quantitative NMR (qNMR), with the resulting calculated concentrations shown in Figure 3C. The methodology is described in Materials and Methods. We found that there was sufficient material remaining in the seized e-cigarettes for quantification for 54 of the 89 positive samples from R1 and R2, with 8 of those samples being at a level below the accurate detection limit of our qNMR methodology (< ∼50 μg mL^-1^). Similarly, we found for these samples that clear peaks were not observable on the LC-MS chromatograms at the dilutions used. We have therefore labelled these samples as ‘low’ in Tables S1-S2.

Combined, the samples have an average concentration of 1.03 ± 0.86 (standard deviation, SD) mg mL^-1^. The average from R1 is 1.13 ± 0.91 mg mL^-1^ and from R2 is 0.92 ± 0.81 mg mL^-1^. From Figure 3C, there is a broad distribution of concentrations, with a maximum of 3.6 mg mL^-1^. The data cannot be fit with a single distribution function, but instead apparently are composed of ‘low’, ‘medium’ and ‘high’ distribution within the broader data set. With only 54 samples, we are not able to accurately model a trimodal distribution with any degree of certainty and so Figure 3C shows a distribution at the mean ± half a standard deviation. The remaining two distributions arise from the remaining ‘low’ / ‘high’ data.

Our data suggests that the SC positive e-cigarettes are relatively evenly distributed between refillable devices and unlabelled bottles. We were interested if the low-high distributions in our concentration data reflected a particular sample type. Figure 3C shows the overlay of the refillable devices, bottles and single use concentration data. These data suggest that the ‘low’ concentration range is dominated by the refillable and single use e-cigarettes (79 % of samples). Conversely the ‘high’ concentration range is dominated by bottles of liquid (81 %). The average SC concentration for bottles of liquid is 1.45 mg ml^-1^ and for refillable devices is 0.75 mg ml^-1^ and noting 28 % of the refillable devices had concentrations below our limit of detection. We would posit that the concentrations are rather lower from the refillable devices potentially due to dilution of a historic SC sample with a non-SC-liquid and this would seem logical. Given almost all SC e-cigarette liquids are sold online as bottles of liquid,^15^ the concentration present in the bottles alone would seem the ‘intended’ concentration.

The concentrations from single use e-cigarettes were extremely low compared to liquids and refillable device, being 0.20 ± 0.14 mg mL^-1^. We cannot confidently identify how the SC e-liquid entered these single use e-cigarettes. Whilst it is technically non-trivial to take apart an e-cigarette and add to / replace the liquid in the internal sponge, it is not impossible. Alternatively, SC e-liquid could be added dropwise through the mouthpiece, though we have low confidence in what this would mean for a person using the e-cigarette in terms of dosage. In any case, that these single use e-cigarettes are both relatively rare and at a ‘low’ concentration suggests that at present these are not the critical modality of concern.

Figures 3D and 3E show the correlation between the percentage of SC e-cigarette presence *versus* social metrics from Table 1. We have included only those schools where there are > 20 submitted samples in an effort to decrease sampling bias, and this includes 4 schools from R1, 5 schools from R2 and 2 from R3. The data represents 82 % of all the seized e-cigarettes reported in this study. From Figure 3D-E, there is potentially a positive (linear) correlation between the presence of SC e-cigarettes and free school meals, persistent absence and fraction of SEN support. From a Pearson’s correlation analysis, we find that the fraction of free school meals (FSM) is positively correlated, r = 0.65 and p = 0.01, respectively. However, we find weak/no correlation with persistent absence (r = 0.30, p = 0.002) (, the percentage of SEN support (r = 0.67, p = 0.85) and EHCPs (r = 0.12, p = 0.007). We accept the relatively small samples size (11 schools) used to establish the correlations. However, the statistics suggest an underlying trend with a metric of social deprivation, the fraction of free schools meals.

As a final sampling effort, we have sampled a region (R4) that is geographically distinct from R1-R3, but where the samples are identified by GC-MS. These samples are biased in that they represent samples that were considered suspicious by the police force in R4. Fifty samples were collected over the period November 2023 – July 2024 from four different schools in R4. Because of the nature of the sampling of R4, we do not include these data in the percentage of SC e-cigarettes at the school level. However, these data are particularly interesting because they report the specific SC identified and the physical presentation. We therefore treat these data as a separate validation of the general findings from R1-R3 and to establish whether the findings are borne out when a random region of interest is selected. These data are given in Table S5 and photographs of the positive samples are given in Figure S10. In terms of physical presentation, these samples track with our findings in Figure 3B, with 75 % of SC positives being in refillable devices, 19 % in unlabelled bottles, 6 % in single use e-cigarettes (1 example) and none in labelled bottles. 75 % of samples contained MDMB-4en-PINACA (compared to 93 % in R1-R2) and 25 % containing 4F-MDMB-BINACA. Finally, two THC single use e-cigarettes were identified.

Similar to the findings in R1-3, one of these is a commercially available product, that appears to have been imported and the other is a device marketed for THC oil/resin and having the same form factor as those shown in Figure S9. That is, the data from R4 mirror our findings from R1-3 and increase the extent to which our findings may be generalisable to other schools in England.

## Discussion

In the first study to identify and quantify SCs in e-cigarettes routinely collected from schools, we report an alarmingly high prevalence (17.4 %) of e-cigarettes containing SCs in English schools. While the generalisability of this estimate should be interpreted with caution due to the non-probability sampling methodology and sample size, we note that R1 – R4 represent a very large North-South length span of England (250 miles), consisting of five different regions in the UK, all of which are at least 90 miles apart. That is, these results should raise caution for schools across the country.

The findings of SC e-cigarette presence are directly contrasted by the lack of THC e-cigarettes. This is surprising given that 31 % of people under 17 report having used cannabis^23^ and 6-8% of secondary school students 11-15 years of age report having used cannabis in the past year^24^ Indeed, since the 1980s, the social view of cannabis use is that it is increasingly an unremarkable feature of adolescent life.^25^

How can these findings be reconciled? Our hypothesis is that young people are unaware that the e-cigarettes they buy are SC e-cigarettes and are instead sold under the impression they are ‘cannabis’. The evidence from WEDINOS (above)^16^ support this notion, with SC e-cigarettes almost never being the purchase intent of samples submitted to this drug checking service. Our data argues that *bone fide* THC e-cigarettes are relatively rare compared to SC e-cigarettes. Where THC e-cigarettes are present, they are typically a commercial product from a country where THC is legal and with a relatively high price point (>∼ £15). This compares with as little as ∼£3 for 2 mL (a full e-cigarette) of SC liquid. The considerably lower price of SC compared with THC e-cigarettes together with an assumed inability of consumers to determine their content, may explain the high prevalence of SCs in e-cigarettes. Moreover, we would argue this trend is relatively recent, given the media reporting on the issue appears to have essentially ‘peaked’ in the UK in the year 2023/2024 (the time of writing).

The major SC identified was MDMB-4en-PINACA in all regions where we have identification. However, in total we have identified six different SCs, which highlights the need for generic detection capability as the availability and prevalence of different SCs changes.

### Identification of SC e-cigarettes

Our data highlight that SC e-cigarettes are easy to identify by eye, with nearly half (Table 1, 47.4 %) of both unlabelled bottles and refillable devices containing SCs. This tracks with findings that the available modality from web shops is vastly via bottles of liquid (99.6 % of products).^17^ THC e-cigarettes and the refillable devices used for SC liquid have a different form factor, with the former being specifically designed for THC oil/resin. We accept that the distinction would require specific training using exemplar material, but we feel this would be possible. That is, a teacher or other responsible adult could, as a reasonable guess, establish the high probability of SCs / THC being present in refillable devices and unlabelled bottles, and particularly when combining information from the colour of the liquid (blue, green, red, pink, purple, essentially always being SC e-cigarette liquid). This visual triage could then be useful for identifying samples of concern to the police for further investigation and decrease the sample volume burden for analysis.

### The presence of SC e-cigarettes tracks with a metric of social deprivation

Our data shows a positive correlation with a specific metric of social deprivation; the fraction of free school meals. This further highlights the potential importance of the low price of SCs when compared to THC in e-cigarettes, as discussed above. Whilst we acknowledge the weaknesses in a sample size of 11 schools, we point to the distribution of the schools across R1-R3 and that we have selected them based on a meaningful cut-off of sample size (20 submissions minimum). There is a risk when discussing such data that certain groups become stigmatised, or otherwise adversely affected by the findings. Indeed, only a single school in Figure 3D and 3E had no incidence of SC e-cigarettes. We would point to the ample evidence that the *harm* arising from drugs becomes greater with metrics of social deprivation,^26^ for example becoming involved in county lines drug supply. That is, prior to this article, SCs were considered to almost entirely be found in prisons and in homeless communities, essentially a drug used by the most vulnerable and marginalised. We would suggest that this new evidence less reflects a link between the usage of SC-cigarettes based on social factors and more on the heightened risk that children who use SC e-cigarettes are subject too. Any policy considerations that arise from this work should have this factor at the core of any response.

## Conclusions

Herein, we have demonstrated the consumption of SCs via e-cigarette in schools across England. They are almost exclusively supplied as a liquid refill to a reusable e-cigarette pen. The presence of SC e-cigarettes is counterpointed by the lack of THC e-cigarettes, and we suggest this is because pupils are being sold SC e-cigarettes as ‘cannabis’. Indeed, online webstores market SCs as essentially interchangeable with cannabis, imply it is legal and the cost is many times lower than for THC.^17^ Young people consider cannabis as relatively safe to consume,^22^ and e-cigarettes are prevalent in schools.^14^ That is, there appear to be a confluence of factors that may drive SC e-cigarette use by young people. Crucially, there are no studies examining the pharmacology of SCs in children and so the true risk to health in both the immediate and long term is unknown. Moreover, if young people are not aware they are using / have used SCs, this limits the ability of healthcare providers to provide appropriate support.

Our findings have immediate implications for harm reduction interventions that should be developed for schools, as well as the stark need to better understand the risk to the health of children when they consume SCs, both acute and longer term. There is also a need to expand this work to inform the targeting of intervention and establish whether the prevalence of SCs in e-cigarettes is high across the wider UK region, or if this phenomenon is geographically or socially patterned.

Finally, our findings are immediately applicable to policy, demonstrating that banning single use e-cigarettes will not meaningfully affect the presence of SCs in schools, precisely because they are relatively rare in single use e-cigarettes. An unintended consequence could be an increase in the use of refillable devices and the potential exposure to SCs within e-cigarette liquids. This intelligence is further useful for policing efforts in the community and to identify modes of supply. We highlight that education can be an effective tool in combating drug-related harm, particularly where young people are involved and well informed.

## Materials and Methods

### Sample acquisition

For R1-R3 we initially engaged with the regional police force to request local schools to submit ecigarettes that had been confiscated on the school premises by teachers. No specific schools were targeted for submissions, rather the local force area requested schools to submit samples. The schools were not given any instruction or intelligence on the kind of e-cigarettes to include in their submission. The regional force then collated all submitted samples without a further selection step. For R1-R2 the relevant police force conveyed the samples to our laboratories. For R3, we collected data from samples at a local police station in R3 using our portable technology described above. We note that any positives were immediately handed to a police officer who continuously chaperoned the testing. Sampling for R4 was biased, in that local police officers self-identified samples of concern, or the samples were selected based on another bias e.g. presentation of the material. We note the data reporting on the fraction of SC e-cigarettes (Table 1) arises solely from the unbiased sampling in R1-3.

R1-3 were identified after an initial interaction with the local police force to seek permission to sample. Our rationale for contacting R1-3 was the occurrence of a media report of SC intoxication, relevant to that police forces jurisdiction. We note that we have contact other police forces who did not wish to participate during the time period of the study, or where the local council was not willing to allow the sampling to proceed. As noted in the text R4 arises from biased sampling where schools/police forces requested we identify specific samples, or there was another triage step applied to the samples, either subjective or objective.

### LC-MS

For LC-QToF-MS analysis, all e-cigarette liquid samples were initially diluted 20 000x in HPLC grade ethanol, with repeats at higher concentration where needed/possible. Analyses were performed using an Agilent QToF 6545 with a Jetstream electrospray ionization (ESI) source coupled to an Agilent 1260 Infinity II Quaternary pump HPLC with a 1260 autosampler, column oven compartment, and variable wavelength detector (VWD). The mobile phases used were (A) LC-MS grade water with 0.1% formic acid and (B) acetonitrile with 0.1% formic acid. The gradient used was 95:5 A:B from 0.00-0.60 min, change to 0:100 A:B over 0.60-3.00 min, held at 0:100 A:B from 3.00-5.50 min, change to 95:5 A:B over 5.50-5.60 min, and held at 95:5 A:B from 5.6-7.6 min. The flow rate was 0.5 mL/min at 50ºC and 5 μL of the sample was injected onto an EC-C18 3.0 × 50 mm, 2.7 μm particle size column (InfinityLab Poroshell 120, Agilent Technologies). The MS was operated in positive ionization mode with the gas temperature at 250°C, the drying gas at 11 L/min, and the nebulizer gas at 35 psi (2.41 bar). The sheath gas temperature was set to 300ºC and the flow rate was 12 L/min. The MS was calibrated using a reference calibrant introduced from an independent ESI reference sprayer. The VCap, Fragmentor and Skimmer were set to 3500, 160, and 45 V, respectively. The MS was operated in all-ions mode with three collision energy scan segments at 0, 20, and 40 eV.

The VWD was set to detect at 280 nm wavelength at a frequency of 2.5 Hz. Data processing was automated in Qual 10, with the molecular feature extraction set to the largest 20 compounds for [M^+^H]^+^, [M-H]^-^, and [M+HCOO]^-^ ions.

The results were also searched against the online mass spectral databases HighResNPS (containing over 2300 unique compound entries) and ForTox, with a forward score of 25 and reverse score of 70, and mass tolerances within 5 ppm of the reference library matches. Qualified ions had co-elution scores of ≥ 90, retention time tolerances of ± 0.10, and a minimum S/N of ≥ 5.00.

### GC-MS

Samples were diluted with methanol 50/50 ratio. GCMS analysis was preformed using a TQ-GC (Waters Corporation). Chromatographic separation was performed on a Thames Restek (DB-5, 30 m x 0.25 mm ID x 0.25 μm).

The following temperature parameters were used: 60 °C (held for 3min) to 300 °C at 15 °C min-1, then held for 6 min. Injection port was 280°C, split ratio 25 mL/min. The analyser was set to scan m/z 50-500. GCMS peaks were identified through comparison to standards, NIST database and Cayman Chemicals forensic database.

### NMR

The quantitative nuclear magnetic resonance (NMR) method used was based on a study that quantified SCs in seized e-liquids.^18^ Samples were prepared by mixing 200 μL of e-cigarette liquid with 400 μL of methanol-d4 (MeOD) containing 3 mg of 3-(trimethylsilyl)propionic-2,2,3,3-d4 acid sodium salt (purity ≥ 99%, isotopic purity 98 atom % D) (TSP). A set of MDMB-4en-PINACA concentrations (10 – 0.1 mg/ml) were prepared in 50/50 polyethylene glycol/glycerol (PG/VG) as standard e-cigarette liquids to test the quantification method.

^1^H NMR data were recorded on a Bruker AvanceCore 400 MHz spectrometer (1H frequency of 400.130), with a zg pulse sequence composed of 3.18 s acquisition time, 128 scans and 20 s delay. Chemical shifts were referenced to 3.31 ppm for residual CD2HOD solvent peak (from MeOD) and are reported in ppm. NMR spectra were processed with Mestralab Mnova 14.1 using automatic phase and Whittaker smoother baseline corrections, followed by zero filling (4 x original size) and line broadening (1 Hz) to improve signal/noise ratio. Due to the large amounts of PG/VG in e-cigarette liquid, only the 4 peaks from the aromatic indazole core of the SCs could be reliably integrated. As many of these were used for the qNMR calculation, when not obscured by other additives in the e-cigarette liquid.

The following equation was used for the ^1^H qNMR quantitation:

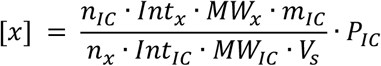

Where [] denotes concentration in mg/mL, P is the purity, n is the number of protons, Int is the integral value, MW is the molecular weight, m is the mass in mg, V is the volume in mL, IC is the internal calibrant, x is the analyte, and s is the sample. As the indazole peaks of the different SC compounds overlayed, when multiple SC compounds were present in each sample, the molecular weight of the main SC, as judged by the LC-MS chromatogram, was used in the calculation.

## Data Availability

All data produced in the present study are available upon reasonable request to the authors

## ASSOCIATED CONTENT

### Supporting Information

Tabulated data from LC-MS, GC-MS and NMR data. Tables giving data for individual samples. Photographs of specific samples noted in the main text.

## AUTHOR INFORMATION

### Author Contributions

The manuscript was written through contributions of all authors. All authors have given approval to the final version of the manuscript.

## ACKNOWLEDGMENT

We gratefully acknowledge the support of a range of anonymous police forces and schools across England.

## ABBREVIATIONS

SC: synthetic cannabinoid
THC: tetrahydrocannabinol
LC/GC-MS: liquid/gas chromatography-mass spectrometry
NMR: nuclear magnetic resonance

## REFERENCES

1 European Monitoring Centre for Drugs and Drug Addiction. New Psychoactive Substances: 25 Years of Early Warning and Response in Europe; EMCDDA: Lisbon, 2022.

2 Andrews, R.; Jorge, R.; Christie, R.; Gallegos, A. From JWH-018 to OXIZIDS: Structural Evolution of Synthetic Cannabinoids in the European Union from 2008 to Present Day. Drug Test. Anal. 2023, 15 (4), 378–387.

3 Hindson, S. A.; Andrews, R. C.; Danson, M. J.; van der Kamp, M. W.; Manley, A. E.; Sutcliffe, O. B.; Haines, T. S. F.; Freeman, T. P.; Scott, J.; Husbands, S. M.; Blagbrough, I. S.; Anderson, J. L. R.; Carbery, D. R.; Pudney, C. R. Synthetic Cannabinoid Receptor Agonists Are Monoamine Oxidase-A Selective Inhibitors. FEBS J. 2023, 290 (12), 3243–3257.

4 Kong, T. Y.; Kim, J. H.; Kim, D. K.; Lee, H. S. Synthetic Cannabinoids Are Substrates and Inhibitors of Multiple Drug-Metabolizing Enzymes. Arch. Pharm. Res. 2018, 41 (7), 691–710.

5 Zagzoog, A.; Brandt, A. L.; Black, T.; Kim, E. D.; Burkart, R.; Patel, M.; Jin, Z.; Nikolaeva, M.; Laprairie, R. B. Assessment of Select Synthetic Cannabinoid Receptor Agonist Bias and Selectivity between the Type 1 and Type 2 Cannabinoid Receptor. Sci. Rep. 2021, 11 (1), 10611.

6 Office for National Statistics. Crime in England and Wales: Year Ending March 2024. https://www.ons.gov.uk/peoplepopulationandcommunity/crimeandjustice/bulletins/crimeinenglandandwales/yearendingmarch 2024 (accessed 2024).

7 Corazza, O.; Coloccini, S.; Marrinan, S.; Vigar, M.; Watkins, C.; Zene, C.; Negri, A.; Aresti, A.; Darke, S.; Rinaldi, R.; Metastasio, A.; Bersani, G. Novel Psychoactive Substances in Custodial Settings: A Mixed Method Investigation on the Experiences of People in Prison and Professionals Working with Them. Front. Psychiatry 2020, 11, 460.

8 Ralphs, R.; Williams, L.; Askew, R.; Norton, A. Adding Spice to the Porridge: The Development of a Synthetic Cannabinoid Market in an English Prison. Int. J. Drug Policy 2017, 40, 57–69.

9 Gray, P.; Ralphs, R.; Williams, L. The Use of Synthetic Cannabinoid Receptor Agonists (SCRAs) within the Homeless Population: Motivations, Harms and the Implications for Developing an Appropriate Response. Addict. Res. Theory 2021, 29, 1–10.

10 Duke, K.; Gleeson, H.; MacGregor, S.; Thom, B. The Risk Matrix: Drug-Related Deaths in Prisons in England and Wales, 2015–2020. J. Community Psychol. 2023, 1–22.

11 Abdulrahim, D.; Bowden-Jones, O. Harms of Synthetic Cannabinoid Receptor Agonists (SCRAs) and Their Management; London, UK, 2016.

12 Craft, S., Ferris, J. A., Barratt, M. J., Maier, L. J., Lynskey, M. T., Winstock, A. R., & Freeman, T. P. (2022). Clinical withdrawal symptom profile of synthetic cannabinoid receptor agonists and comparison of effects with high potency cannabis. Psychopharmacology, 1–9.

13 Norman, C. A Global Review of Prison Drug Smuggling Routes and Trends in the Usage of Drugs in Prisons. WIREs Forensic Sci. 2023, 5 (2), e1473.

14 Craft, S.; Austin, A.; Tooth, T.; Glover, C.; Garrett, T.; Blagbrough, I. S.; Sunderland, P.; Pudney, C. R.; Freeman, T. P. Synthetic Cannabinoid Use in an Adult Male Prison in the UK. Int. J. Drug Policy 2023, 122, 104219. 10.1016/j.drugpo.2023.104219.

15 NHS Digital. Smoking, Drinking and Drug Use among Young People in England, 2021. https://digital.nhs.uk/data-and-information/publications/statistical/smoking-drinking-and-drug-use-among-young-people-in-england/2021/introduction (accessed 2024).

16 Webtool available at: https://www.wedinos.org/ (accessed 2024).

17 Gould, A.; Dargan, P. I.; Wood, D. M. An Internet Snapshot Survey Assessing the Sale of Synthetic Cannabinoid Receptor Agonists for Use with Electronic Vaping Devices. J. Med. Toxicol. 2024, 20, 271–277.

18 Wu, N.; Danoun, S.; Balayssac, S.; Malet-Martino, M.; Lamoureux, C.; Gilard, V. Synthetic Cannabinoids in E-Liquids: A Proton and Fluorine NMR Analysis from a Conventional Spectrometer to a Compact One. Forensic Sci. Int. 2021, 324, 110813. 10.1016/j.forsciint.2021.110813.

19 Winstock, A., Lynskey, M., Borschmann, R., & Waldron, J. (2015). Risk of emergency medical treatment following consumption of cannabis or synthetic cannabinoids in a large global sample. Journal of Psychopharmacology, 29(6), 698–703.

20 Webtool available at: https://www.gov.uk/school-performance-tables?_ga=2.185757866.2019012831.1720090767-359797626.1720090767 (accessed 2024).

21 Cozier, et al. Instant Detection of Synthetic Cannabinoids on Physical Matrices, Implemented on a Low-Cost, Ultraportable Device. Anal. Chem. 2023, 95, 13829.

22 Gardner M, Bowden C, Manzoor S, Cozier G, Andrews R, Craft S, et al. Automated detection of controlled substances from sealed e-cigarettes. ChemRxiv. 2024; doi:10.26434/chemrxiv-2024-ns335

23 Fitzsimons, E.; Villadsen, A. Substance Use and Antisocial Behaviour in Adolescence. https://cls.ucl.ac.uk/wp-content/uploads/2017/02/CLS-briefing-paper-Risky-behaviours-MCS-Age-17-initial-findings.pdf (accessed 2024).

24 NHS Digital. Smoking, Drinking and Drug Use among Young People in England, 2021. https://digital.nhs.uk/data-and-information/publications/statistical/smoking-drinking-and-drug-use-among-young-people-in-england/2021 (accessed 2024).

25 Aldridge, J.; Measham, F.; Williams, L. Illegal Leisure Revisited: Changing Patterns of Alcohol and Drug Use in Adolescents and Young Adults; Routledge: London, 2011; ISBN 978-0415495530.

26 Stevens, A. Drugs, Crime and Public Health: The Political Economy of Drug Policy; Routledge: London, 2011; ISBN 978-0415610674

