## Supporting information for "Synthetic cannabinoids consumed via e-cigarettes in English schools"

### *Supporting information* - Synthetic cannabinoids consumed via e-cigarettes in English schools

---

Table S1. Results summary for all samples in R1

Table S2. Results summary for all samples in R2

Table S3. Results summary for all samples in R3

Table S4. Data summary for R1-3

Table S5. Results summary for all samples in R4

Table S6. LC-MS parent molecule and fragment mass to charge (mz) ratios used to confirm compounds found in the e-cigarettes/liquids

Figure S1. Example LC-MS chromatogram and spectra for sample R1S1.2 with MDMB-4en-PINACA, 4F-MDMB-BINACA and MDMB-INACA SCs identified

Figure S2. Example LC-MS chromatogram and spectra for sample R1S12.119 with ADB-BUTINACA identified

Figure S3. Structures of illicit drugs identified

Figure S4. Example 1H NMR spectra used for qNMR on sample R1S8.62 containing MDMB-4en-PINACA

Figure S5. Example 1H NMR spectra used for qNMR on sample R2S1.3 containing MDMB-4en-PINACA

Figure S6. Example 1H NMR spectra used for qNMR on sample R1S12.121 containing ADB-BUTINACA

Figure S7. Example 1H NMR spectra used for qNMR on sample R1S2.13 containing MDMB-4en-PINACA

Figure S8. Plot showing the qNMR calculated values of a standard concentration range of MDMB-4en-PINACA in e-cigarette liquid

Figure S9. Photographs of THC e-cigarettes from R1-3

Figure S10. Photographs of SC and THC e-cigarettes from R4

**Table S1. Results summary for all samples in R1**

| R1S1 | Sample type | Detail | Liquid colour | LC-MS result | Quant (mg / mL) |
|---|---|---|---|---|---|
| 1 | RF | Vaporesso | Orange | NDD | - |
| 2 | RF | Vaporesso | Yellow | MDMB-4en-PINACA<br>4F-MDMB-BINACA<br>MDMB-INACA | 0.63 |
| 3 | LB | 'Elux Mr Blue', Yellow Liquid | Yellow | NDD | - |
| 4 | LB | 'Elux Lemon and Lime', Yellow Liquid | Yellow | NDD | - |
| 5 | RF | - | - | MDMB-4en-PINACA | IM |
| 6 | RF | Vaporesso | Brown | NDD | - |
| 7 | RF | Vaporesso | Green | MDMB-4en-PINACA<br>4F-MDMB-BINACA<br>MDMB-INACA | 0.14 |
| 8 | RF | Vaporesso | Yellow | MDMB-4en-PINACA<br>4F-MDMB-BINACA<br>MDMB-INACA | IM |
| 9 | UB | - | Yellow | MDMB-4en-PINACA<br>4F-MDMB-BINACA<br>MDMB-INACA | 0.89 |
| R1S2 | Sample type | Detail | Liquid colour | LC-MS result | Quant (mg / mL) |
| 10 | RF | Vaporesso | Clear | MDMB-4en-PINACA<br>MDMB-INACA | 0.11 |
| 11 | SU | - | - | NT | - |
| 12 | SU | Gold bar | Yellow | NDD | - |
| 13 | LB | 'Elux Mr Blue', Yellow Liquid | Blue | MDMB-4en-PINACA<br>MDMB-PINACA | 2.69 |
| 14 | RF | Vaporesso | Green | MDMB-4en-PINACA<br>MDMB-INACA | 1.32 |
| R1S3 | Sample type | Detail | Liquid colour | LC-MS result | Quant (mg / mL) |
| 15 | SU | Crystal bar, Cherry fizz | - | NT | - |
| R1S4 | Sample type | Detail | Liquid colour | LC-MS result | Quant (mg / mL) |
| 16 | SU | Enjoy Ultra | - | NT | - |
| 17 | SU | OXAV | - | NT | - |
| 18 | RF | Vaporesso, orange liquid | Yellow | MDMB-4en-PINACA<br>MDMB-INACA<br>4F-MDMB-BINACA | 0.56 |
| 19 | SU | Hit Fusion, Pink Lemonade | - | NT | - |
| 20 | SU | Elf Bar | - | NDD | - |
| R1S5 | Sample type | Detail | Liquid colour | LC-MS result | Quant (mg / mL) |
| 21 | SU | Crystal Bar, Pineapple peach mango | - | NT | - |
| 22 | SU | Crystal Bar, Cola ice | - | NT | - |
| 23 | SU | Crystal bar, Cola ice | - | NT | - |
| R1S6 | Sample type | Detail | Liquid colour | LC-MS result | Quant (mg / mL) |
| 24 | SU | Randm Tornado | - | NT | - |
| 25 | SU | Sky Hunter | - | NT | - |
| 26 | RF | Vaporesso | Brown | MDMB-4en-PINACA | 0.29 |
| 27 | UB | - | Yellow | MDMB-4en-PINACA | 0.51 |

| R1S7 | Sample type | Detail | Liquid colour | LC-MS result | Quant (mg / mL) |
|---|---|---|---|---|---|
| | | | | 4F-MDMB-BINACA | |
| 28 | SU | Lost Mary, Blue Razz Cherry | - | NDD | - |
| 29 | SU | Crystal Bar, Blueberry sour raspberry | - | NT | - |
| 30 | SU | Crystal Bar, Blueberry peach ice | - | NDD | - |
| 31 | SU | Twister bar, Strawberry raspberry cherry | - | NDD | - |
| 32 | SU | Elfbar | - | NDD | - |
| 33 | SU | Lost Mary, Pineapple ice | - | NDD | - |
| 34 | SU | Lost Mary, Cherry ice | - | NT | - |
| 35 | SU | Lost Mary, Cherry ice | - | NDD | - |
| 36 | SU | Elfbar, Cola | - | NDD | - |
| 37 | SU | Brand unknown | - | NT | - |
| 38 | SU | Crystal Bar, Lemon and Lime | - | NDD | - |
| 39 | SU | Elfbar | - | NDD | - |
| 40 | SU | Lost Mary, Pineapple ice | - | NT | - |
| 41 | SU | Crystal Bar, Tiger blood | - | NDD | - |
| 42 | SU | Crystal bar, Lemon and Lime | - | NDD | - |
| 43 | SU | Lost Mary, Pineapple ice | - | NDD | - |
| 44 | SU | Elfbar | - | NDD | - |
| 45 | SU | Lost Mary, Double Apple | - | NT | - |
| 46 | SU | Lost Mary, Blue Razz ice | - | NT | - |
| 47 | SU | Lost Mary, Kiwi passion fruit guava | - | NDD | - |
| 48 | SU | Bloody Mary Crystal, Strawberry ice | - | NT | - |
| 49 | SU | Crystal Bar, Lemon and Lime | - | NT | - |
| 50 | RF | Vaporesso | Yellow | MDMB-4en-PINACA | IM |
| 51 | SU | Crystal Bar, Lemon and Lime | - | NT | - |
| 52 | RF | Vaporesso | Green | ADB-BUTINACA<br>MDMB-4en-PINACA | 2.04 |
| 53 | RF | Vaporesso | Yellow | MDMB-4en-PINACA | 'low' |

| R1S8 | Sample type | Detail | Liquid colour | LC-MS result | Quant (mg / mL) |
|---|---|---|---|---|---|
| 54 | SU | Unknown brand, Cherry ice | Yellow | NDD | - |
| 55 | LB | Labelled bottle, Elux, Blueberry raspberry, yellow liquid | Yellow | NDD | - |
| 56 | RF | Vaporesso, yellow liquid | Yellow | MDMB-4en-PINACA | 'low' |
| 57 | RF | Vaporesso, yellow liquid | Yellow | NDD | - |
| 58 | RF | Vaporesso, Brown liquid | Brown | NDD | - |
| 59 | LB | Labelled bottle, Frunk, Pineapple express, Clear liquid | Clear | NDD | - |
| 60 | LB | Labelled bottle, Elux, MR Blue, Green liquid | Green | MDMB-4en-PINACA | 'low' |
| 61 | LB | Labelled bottle, Elux, Cherry ice, yellow liquid | Yellow | NDD | - |
| 62 | UB | - | Clear | MDMB-4en-PINACA | 3.52 |
| 63 | RF | Vaporesso, orange liquid | Yellow | MDMB-4en-PINACA | 'low' |
| 64 | LB | Labelled bottle, Elux, Blueberry cherry cranberry, orange liquid | Yellow | MDMB-4en-PINACA | 'low' |
| 65 | RF | Vaporesso, green liquid | Green | MDMB-4en-PINACA | 1.31 |
| 66 | SU | Crystal bar, Cherry ice | | NT | - |
| 67 | SU | Crystal bar, Watermelon strawberry | - | NT | - |

| | | | | | |
|---|---|---|---|---|---|
| 68 | SU | Crystal bar, Lemon and Lime | - | NT | - |
| 69 | SU | Crystal bar, Cherry ice | Yellow | NDD | - |
| 70 | RF | Vaporesso, yellow liquid | Yellow | MDMB-4en-PINACA | 0.37 |
| 71 | RF | Vaporesso, green liquid | Green | MDMB-4en-PINACA<br>ADB-4en-PINACA | 0.39 |
| 72 | SU | Randm tornado | Yellow | MDMB-4en-PINACA | IM |
| 73 | RF | Vaporesso, green liquid | Green | NDD | - |
| 74 | SU | R and M, Magic mint | - | NDD | - |
| 75 | RF | Arc Mini | - | NT | - |
| 76 | SU | Vuse Go | - | NT | - |
| 77 | SU | Crystal Bar, Sour apple blueberry | - | NDD | - |
| 78 | SU | Crystal Bar, Fizzy cherry | - | NDD | - |
| 79 | RF | Vaporesso | Yellow | MDMB-4en-PINACA | ‘low’ |
| 80 | SU | Bloody Mary, triple berry | Yellow | NDD | - |
| 81 | SU | Crystal bar, Lemon and Lime | - | NT | - |
| 82 | SU | Crystal bar, Lemon and Lime | Yellow | NDD | - |
| 83 | LB | Labelled bottle, Elflic, strawberry ice cream, yellow liquid | Yellow | NDD | - |
| 84 | SU | Elfbar, Strawberry raspberry cherry ice | Yellow | NDD | - |
| 85 | UB | Unlabelled bottle, clear liquid | Clear | NDD | - |
| 86 | SU | Zillion | Yellow | NDD | - |
| 87 | SU | Elfbar, watermelon | Yellow | NDD | - |
| 88 | LB | Zeus, clear liquid | Clear | NDD | - |
| 89 | LB | Zeus, clear liquid | Clear | NDD | - |
| R1S9 | Sample type | Detail | Liquid colour | LC-MS result | Quant (mg / mL) |
| 90 | RF | Vaporesso | Green | MDMB-4en-PINACA | 0.42 |
| 91 | RF | Vaporesso | Yellow | NDD | - |
| 92 | RF | Vaporesso | Green | MDMB-4en-PINACA | 0.36 |
| 93 | UB | - | Blue | MDMB-4en-PINACA | 2.6 |
| 94 | UB | - | Blue | MDMB-4en-PINACA<br>MDMB-PINACA | 1.24 |
| 95 | SU | Enjoy Ultra, peach watermelon strawberry | - | NDD | - |
| R1S10 | Sample type | Detail | Liquid colour | LC-MS result | Quant (mg / mL) |
| 96 | SU | Crystal bar | Yellow | NDD | - |
| 97 | SU | Elfbar | Yellow | NDD | - |
| 98 | SU | Gold bar | Yellow | NDD | - |
| 99 | SU | Crystal bar | Yellow | NDD | - |
| 100 | SU | Crystal bar | Yellow | NDD | - |
| 101 | SU | Ene Legend | Yellow | NT | - |
| 102 | SU | Hyati pro | Yellow | NDD | - |
| 103 | SU | Elux legend pro | Yellow | NDD | - |
| 104 | SU | Elfbar | Brown | NDD | - |
| 105 | SU | Crystal bar | Yellow | NT | - |
| 106 | SU | Crystal bar | Yellow | NT | - |
| 107 | SU | Elfbar | Brown | NDD | - |
| 108 | SU | Randm Tornado | Yellow | NDD | - |
| 109 | SU | Elfbar | Yellow | NDD | - |

| | | | | | |
|---|---|---|---|---|---|
| 110 | SU | Hyati pro | Yellow | NDD | - |
| 111 | SU | Lost mary | Yellow | NT | - |
| 112 | SU | Gold bar | Yellow | NDD | - |
| 113 | SU | Crystal bar | Yellow | NT | - |
| 114 | SU | - | Yellow | NDD | - |
| 115 | SU | Ene Legend | Brown | NDD | - |
| 116 | SU | - | Brown | NDD | - |
| R1S11 | Sample type | Detail | Liquid colour | LC-MS result | Quant (mg / mL) |
| 117 | UB | - | Pink | MDMB-4en-PINACA<br>MDMB-PINACA | 1.5 |
| R1S12 | Sample type | Detail | Liquid colour | LC-MS result | Quant (mg / mL) |
| 118 | RF | Vaporesso | Red | MDMB-4en-PINACA | 'low' |
| 119 | UB | - | Purple | ADB-BUTINACA | 2.16 |
| 120 | UB | - | Yellow | ADB-BUTINACA | 1.19 |
| 121 | UB | - | Yellow | ADB-BUTINACA | 1.28 |
| 122 | UB | - | Green | MDMB-4en-PINACA<br>4F-MDMB-BINACA<br>MDMB-INACA | 1.28 |
| 123 | UB | - | Green | MDMB-4en-PINACA<br>4F-MDMB-BINACA<br>MDMB-INACA | 0.34 |
| 124 | SU | - | Brown resin | THC | 508.15 |
| 125 | SU | Crystal bar | - | NT | - |
| 126 | SU | Crystal bar | - | NT | - |
| 127 | SU | Enjoy ultra | - | NT | - |
| 128 | SU | Twins maxfel | - | NT | - |
| 129 | SU | Lost mary | - | NT | - |
| 130 | SU | Enjoy ultra | - | NT | - |
| 131 | SU | 88. vapes | - | NT | - |
| 132 | SU | Hayati pro ultra | - | NT | - |
| 133 | SU | Crystal bar | - | NT | - |
| 134 | SU | Randm tornado | - | NT | - |
| 135 | SU | Randm tornado | - | NT | - |
| 136 | SU | Randm tornado | - | NT | - |
| 137 | SU | 88 vapes | - | NT | - |
| 138 | SU | Crystal original | - | NT | - |
| 139 | SU | Lost mary | - | NT | - |
| 140 | SU | Lost mary | - | NT | - |
| 141 | SU | Randm tornado | - | NT | - |
| 142 | SU | Insta bar | - | NT | - |
| 143 | SU | 88. vapes | - | NT | - |
| 144 | SU | - | - | NT | - |
| 145 | SU | - | - | NT | - |
| 146 | SU | - | - | NT | - |
| 147 | SU | - | - | NT | - |
| 148 | SU | - | - | NT | - |
| 149 | SU | - | - | NT | - |

| | | | | | |
|---|---|---|---|---|---|
| 150 | SU | - | - | NT | - |
| R1S13 | Sample type | Detail | Liquid colour | LC-MS result | Quant (mg / mL) |
| 151 | SU | Twister bar | - | NT | - |
| 152 | SU | Crystal bar | Yellow | NDD | - |
| 153 | SU | Crystal bar | Yellow | NDD | - |
| 154 | SU | Crystal bar | - | NT | - |
| 155 | SU | Crystal bar | - | NT | - |
| 156 | SU | - | - | NT | - |

NDD, no drug detected; 'low', concentration below level of NMR measurement; IM, insufficient material; NT, not tested; SU, single use; RF, refillable; UB, unlabelled bottle; LB, labelled bottle.

**Table S2. Results summary for all samples in R2**

| R2S1 | Sample type | Detail | Liquid colour | LC-MS result | Quant (mg / mL) |
|---|---|---|---|---|---|
| 1 | UB | - | Purple | MDMB-4en-PINACA<br>MDMB-PINACA | 1.71 |
| 2 | UB | - | Clear | MDMB-4en-PINACA | IM |
| 3 | UB | - | Purple | MDMB-4en-PINACA<br>MDMB-PINACA | 1.56 |
| 4 | UB | - | Blue | MDMB-4en-PINACA | 2.92 |
| 5 | RF | Vaporesso | Clear | MDMB-4en-PINACA | IM |
| 6 | UB | - | Yellow | MDMB-4en-PINACA | IM |
| 7 | LB | Elux, Mr Blue | Yellow | NDD | - |
| 8a | SU | Hyati (2$^{nd}$ pad) | Green | MDMB-4en-PINACA | 0.17 |
| 8b | SU | Hyati (main pad) | Yellow | MDMB-4en-PINACA | 0.05 |
| 9 | SU | - | Yellow | NDD | - |
| 10 | SU | - | Yellow | NDD | - |
| 11 | SU | - | Yellow | NDD | - |
| 12 | SU | - | Yellow | NDD | - |
| 13 | SU | - | Yellow | NDD | - |
| 14 | SU | - | Yellow | NDD | - |
| 15 | SU | - | Yellow | NDD | - |
| 16 | SU | - | Yellow | NDD | - |
| 17a | SU | Hyati (2$^{nd}$ pad) | Purple | MDMB-4en-PINACA | 0.4 |
| 17b | SU | Hyati (main pad) | Brown | MDMB-4en-PINACA | IM |
| 18 | SU | - | Brown | NDD | - |
| 19 | SU | - | - | NT | - |
| 20 | SU | - | - | NT | - |
| 21 | SU | - | - | NT | - |
| 22 | SU | Randm Tornado | Yellow | MDMB-4en-PINACA<br>MDMB-INACA | IM |
| 23 | SU | Randm tornado | Green | MDMB-4en-PINACA | 0.3 |
| 24 | SU | - | Clear | NDD | - |
| 25 | SU | Quoa | Yellow | MDMB-4en-PINACA | 0.14 |
| 26 | SU | - | Yellow | NDD | - |
| 27 | SU | - | - | NT | - |
| 28 | SU | - | - | NT | - |
| 29 | SU | Hyati pro ultra | Yellow | MDMB-4en-PINACA | 0.15 |
| 30 | SU | - | Yellow | NDD | - |
| 31 | SU | - | Yellow | NDD | - |
| 32 | SU | - | Yellow | NDD | - |
| 33 | SU | - | Yellow | NDD | - |
| R2S2 | Sample type | Detail | Liquid colour | LC-MS result | Quant (mg / mL) |
| 34 | RF | - | - | NT | - |
| 35 | RF | Elfbar | Yellow | NDD | - |
| 36 | UB | - | Pink | Heroin<br>MDMB-4en-PINACA<br>ADB-BUTINACA | 0.1 (heroin),<br>0.28 (SC) |
| 37 | RF | 88 Vapes | Blue | MDMB-4en-PINACA | 0.4 |

| | | | | | |
|---|---|---|---|---|---|
| 38 | RF | Elfbar | Yellow | NDD | - |
| 39 | RF | Caliburn | Clear | NDD | - |
| 40 | LB | IVG salt, Cherry bubblegum breeze | Clear | NDD | - |
| 41 | UB | - | Clear | NDD | - |
| 42 | LB | Bar juice, watermelon | Yellow | NDD | - |
| 43 | LB | Elux | Yellow | NDD | - |
| 44 | RF | Vaporesso | Yellow | NDD | - |
| 45 | RF | RPM | Brown | MDMB-4en-PINACA<br>MDMB-INACA | 'low' |
| 46 | SU | Elux legend pro | Brown | NDD | - |
| 47 | SU | Elux legend pro | Brown | NDD | - |
| 48 | SU | Elux legend pro | Brown | NDD | - |
| 49 | SU | Elux legend pro | Brown | NDD | - |
| 50 | SU | Randm tornado | Brown | NDD | - |
| 51 | SU | Randm tornado | Yellow | NDD | - |
| 52 | SU | Randm tornado | Yellow | NDD | - |
| 53 | SU | Ene legend | Yellow | NDD | - |
| 54 | SU | Hyati pro ultra | Yellow | NDD | - |
| 55 | SU | Ene legend | Yellow | NDD | - |
| 56 | SU | Elfbar | Yellow | NDD | - |
| 57 | SU | Elfbar | Brown | NDD | - |
| 58 | SU | Elfbar | Brown | NDD | - |
| 59 | SU | Elfbar | Brown | NDD | - |
| 60 | SU | Elfbar | Yellow | NDD | - |
| 61 | SU | - | Brown | NDD | - |
| 62 | SU | Hyati | Yellow | NDD | - |
| 63 | SU | Hyati pro ultra | Yellow | NDD | - |
| 64 | SU | Lost Mary | Yellow | NDD | - |
| 65 | SU | Lost MAry | Brown | NDD | - |
| R2S3 | Sample type | Detail | Liquid colour | LC-MS result | Quant (mg / mL) |
| 66 | UB | - | Yellow | NDD | - |
| 67 | UB | - | Red | MDMB-4en-PINACA<br>MDMB-INACA | 0.54 |
| 68 | UB | - | Clear | MDMB-4en-PINACA | IM |
| 69 | UB | - | Yellow opaque | NDD | - |
| 70 | UB | - | Yellow | MDMB-BUTINACA<br>MDMB-4en-PINACA<br>MDMB-PINACA | 1.12 |
| 71 | UB | - | Yellow | MDMB-BUTINACA<br>MDMB-INACA | 0.97 |
| 72 | UB | - | Clear | MDMB-4en-PINACA<br>MDMB-INACA | 0.38 |
| 73 | UB | - | Clear | MDMB-4en-PINACA | IM |
| 74 | UB | - | Clear | MDMB-4en-PINACA | IM |
| 75 | RF | 88 Vapes | - | NT | - |
| 76 | RF | 88 Vapes | Yellow | MDMB-4en-PINACA | 'low' |
| 77 | SU | | | NT | - |

| | | | | | |
|---|---|---|---|---|---|
| 78 | RF | Vaporesso | Yellow | MDMB-4en-PINACA<br>ADB-4en-PINACA<br>MDMB-INACA | 1.26 |
| 79 | SU | WGA pro | - | NT | - |
| 80 | LB | - | Clear | NDD | - |
| 81 | LB | - | Yellow | NDD | - |
| 82 | LB | - | Yellow | NDD | - |
| 83 | LB | - | Yellow | NDD | - |
| 84 | LB | - | Yellow | NDD | - |
| 85 | LB | - | Yellow | NDD | - |
| 86 | LB | - | Yellow | NDD | - |
| 87 | RF | Vaporesso | Yellow | NDD | - |
| 88 | RF | Solus | Clear | MDMB-4en-PINACA | IM |
| 89 | RF | KRT | - | NDD | - |
| 90 | RF | Caliburn | Clear | NDD | - |
| 91 | RF | OXVA | Brown | MDMB-4en-PINACA<br>MDMB-BUTINACA<br>MDMB-INACA | 0.27 |
| 92 | RF | Vaporesso | Brown | MDMB-4en-PINACA | 'low' |
| 93 | RF | Just | - | NDD | - |
| 94 | RF | - | Brown | NDD | - |
| 95 | SU | Hyati pro | Yellow | NDD | - |
| 96 | SU | Hyati pro | Yellow | NDD | - |
| 97 | SU | Hyati pro | Yellow | NDD | - |
| 98 | SU | Hyati pro | Yellow | NDD | - |
| 99 | SU | Hyati pro | Yellow | NDD | - |
| 100 | SU | Hyati pro | Yellow | NDD | - |
| 101 | SU | Hyati pro | Yellow | NDD | - |
| 102 | SU | Hyati pro | Brown | NDD | - |
| 103 | SU | Hyati pro | Yellow | NDD | - |
| 104 | SU | Hyati pro | Yellow | NDD | - |
| 105 | SU | Hyati pro | Yellow | NDD | - |
| 106 | SU | Hyati pro | Yellow | NDD | - |
| 107 | SU | Hyati pro | Yellow | NDD | - |
| 108 | SU | Hyati pro | Yellow | NDD | - |
| 109 | SU | Hyati pro | Yellow | NDD | - |
| 110 | SU | Hyati pro | Yellow | NDD | - |
| 111 | SU | Hyati pro | Yellow | NDD | - |
| 112 | SU | Hyati pro | Yellow | NDD | - |
| 113 | SU | Hyati pro | Brown | NDD | - |
| 114 | SU | Hyati pro | Yellow | NDD | - |
| 115 | SU | Crystal pro | - | NT | - |
| 116 | SU | Crystal pro | Yellow | NDD | - |
| 117 | SU | Crystal pro | - | NT | - |
| 118 | SU | WGA | Brown | NDD | - |
| 119 | SU | Hyati pro ultra | Clear | NDD | - |
| 120 | SU | Crystal bar | - | NT | - |

| | | | | | |
|---|---|---|---|---|---|
| 121 | SU | Crystal bar | - | NT | - |
| 122 | SU | Crystal bar | Clear | NDD | - |
| 123 | SU | Crystal bar | Yellow | NDD | - |
| 124 | SU | Crystal bar | - | NT | - |
| 125 | SU | Elux legend | Yellow | NDD | - |
| 126 | SU | Elux legend | Brown | NDD | - |
| 127 | SU | Elux legend | Yellow | NDD | - |
| 128 | SU | Elux legend | Yellow | NDD | - |
| 129 | SU | Elux legend | Yellow | NDD | - |
| 130 | SU | Elux legend | Brown | NDD | - |
| 131 | SU | Elux legend | Yellow | NDD | - |
| 132 | SU | Elux legend | Brown | NDD | - |
| 133 | SU | Ene Legend | Yellow | NDD | - |
| 134 | SU | Ene Legend | Yellow | NDD | - |
| 135 | SU | Randm Tornado | Yellow | NDD | - |
| 136 | SU | Randm Tornado | Yellow | NDD | - |
| 137 | SU | Randm Tornado | Yellow | NDD | - |
| 138 | SU | Randm Tornado | Yellow | NDD | - |
| 139 | SU | Randm Tornado | Yellow | NDD | - |
| 140 | SU | Randm Tornado | Brown | NDD | - |
| 141 | SU | Randm Tornado | Brown | NDD | - |
| 142 | SU | Randm Tornado | Brown | NDD | - |
| 143 | SU | Ene Legend | Yellow | NDD | - |
| 144 | SU | Elux legend | Yellow | NDD | - |
| 145 | SU | Elux legend | Yellow | NDD | - |
| 146 | SU | Crystal bar | Yellow | NDD | - |
| 147 | SU | Crystal bar | - | NT | - |
| 148 | SU | Crystal bar | Yellow | NDD | - |
| 149 | SU | Crystal bar | Yellow | NDD | - |
| 150 | SU | Elfbar | Yellow | NDD | - |
| 151 | SU | Elfbar | Yellow | NDD | - |
| 152 | SU | Elfbar | Clear | NDD | - |
| 153 | SU | Elfbar | Yellow | NDD | - |
| 154 | SU | Elfbar | Yellow | NDD | - |
| 155 | SU | Elfbar | Brown | NDD | - |
| 156 | SU | Elfbar | Yellow | NT | - |
| 157 | SU | Elfbar | Yellow | NDD | - |
| 158 | SU | Elfbar | Yellow | NDD | - |
| 159 | SU | Veev | Clear | NDD | - |
| 160 | SU | Lost mary | Yellow | NT | - |
| 161 | SU | - | Yellow | NDD | - |
| 162 | SU | - | Yellow | NDD | - |
| 163 | SU | Geek | Yellow | NDD | - |
| 164 | SU | - | Yellow | NDD | - |

| R2S4 | Sample type | Detail | Liquid colour | LC-MS result | Quant (mg / mL) |
|---|---|---|---|---|---|
| 165 | UB | - | Yellow | MDMB-4en-PINACA | IM |
| 166 | RF | Vaporesso | Yellow | MDMB-4en-PINACA | IM |

| | | | | | |
|---|---|---|---|---|---|
| 167 | - | - | Yellow | NDD | - |
| R2S5 | Sample type | Detail | Liquid colour | LC-MS result | Quant (mg / mL) |
| 168 | RF | Bear+ Aspire | Yellow | NDD | - |
| 169 | LB | - | Yellow | NDD | - |
| 170 | RF | Smok Fortis | - | NDD | - |
| 171 | RF | OXVA | Brown | NDD | - |
| 172 | RF | Tri Box | Yellow | MDMB-4en-PINACA | IM |
| 173 | SU | Hyati pro ultra | Yellow | NDD | - |
| 174 | SU | Hyati pro | Yellow | NDD | - |
| 175 | SU | Hyati pro | Yellow | NDD | - |
| 176 | SU | Randm Tornado | Yellow | NDD | - |
| 177 | SU | Randm Tornado | Yellow | NDD | - |
| 178 | SU | Hyati pro | Yellow | NDD | - |
| 179 | SU | - | Yellow | NDD | - |
| 180 | SU | - | Yellow | NDD | - |
| 181 | SU | Hyati pro ultra | Yellow | NDD | - |
| 182 | SU | - | Yellow | NDD | - |
| 183 | SU | Elux legend pro | Yellow | NDD | - |
| 184 | SU | Crystal bar | Yellow | NDD | - |
| 185 | SU | Elux legend pro | Brown | NDD | - |
| 186 | SU | Crystal bar | Yellow | NDD | - |
| 187 | SU | Crystal bar | Yellow | NDD | - |
| 188 | SU | Crystal bar | Yellow | NDD | - |
| 189 | SU | Crystal bar | - | NT | - |
| 190 | SU | Crystal bar | Yellow | NDD | - |
| 191 | SU | Crystal bar | Yellow | NDD | - |
| 192 | SU | | | NT | - |
| R2S6 | Sample type | Detail | Liquid colour | LC-MS result | Quant (mg / mL) |
| 193 | UB | - | Pink | MDMB-4en-PINACA<br>MDMB-PINACA | 2.45 |
| 194 | RF | Gotek cart | Pink | MDMB-4en-PINACA<br>MDMB-PINACA | 2.08 |
| 195 | RF | Gotek cart | Green | MDMB-4en-PINACA<br>MDMB-PINACA | 1.04 |
| 196 | SU | Hyati pro | Yellow | NDD | - |
| 197 | SU | Hyati pro | Yellow | NDD | - |
| 198 | SU | Elux legend | Brown | NDD | - |
| R2S7 | Sample type | Detail | Liquid colour | LC-MS result | Quant (mg / mL) |
| 199 | LB | Elux Banana ice | Yellow | NDD | - |
| 200 | LB | Elux, gummy bear | Yellow | NDD | - |
| 201 | LB | Elux, gummy bear | Yellow | NDD | - |
| 202 | SU | Randm Tornado | Yellow | NDD | - |
| 203 | SU | Hyati duo mesh | Yellow | NDD | - |
| 204 | SU | - | Yellow | NDD | - |
| 205 | SU | Lost Mary | Yellow | NDD | - |
| R2S8 | Sample type | Detail | Liquid colour | LC-MS result | Quant (mg / mL) |
| 206 | RF | Vaporesso | Yellow | NDD | - |

| | | | | | |
|---|---|---|---|---|---|
| 207 | SU | Randm Tornado | Yellow | NDD | - |
| 208 | SU | Randm Tornado | Clear | NDD | - |
| 209 | SU | Randm Tornado | Yellow | NDD | - |
| 210 | SU | Hyati pro ultra | Yellow | NDD | - |
| 211 | SU | Hyati pro ultra | Yellow | NDD | - |
| 212 | SU | Crystal bar | Yellow | NDD | - |
| 213 | SU | Crystal bar | Yellow | NDD | - |
| 214 | SU | Crystal bar | Yellow | NDD | - |
| 215 | SU | Hyati pro max | Yellow | NDD | - |
| 216 | SU | Crystal bar | Yellow | NDD | - |
| 217 | SU | 88 vapes | Yellow | NDD | - |
| 218 | SU | - | Yellow | NDD | - |

| R2S9 | Sample type | Detail | Liquid colour | LC-MS result | Quant (mg / mL) |
|---|---|---|---|---|---|
| 219 | RF | X-Priv | Yellow | MDMB-4en-PINACA | IM |
| 220 | UB | - | Yellow opaque | MDMB-4en-PINACA<br>MDMB-INACA | 0.82 |
| 221 | RF | - | Brown | MDMB-4en-PINACA<br>MDMB-INACA | 0.55 |
| 222 | LB | Heizen blue | Yellow | NDD | - |
| 223 | LB | EPIQ | Yellow | NDD | - |
| 224 | LB | Sense by vapearts | Clear | NDD | - |
| 225 | LB | Sense by vapearts | Clear | NDD | - |
| 226 | LB | Vape district, Pineapple mango | Clear | NDD | - |
| 227 | RF | Smok | Clear | NDD | - |
| 228 | SU | Randm Tornado | Clear | NDD | - |
| 229 | RF | - | Brown | 4F-MDMB-BINACA | IM |
| 230 | RF | - | Green | MDMB-4en-PINACA<br>4F-MDMB-BINACA | IM |
| 231 | RF | - | Brown | NDD | - |
| 232 | SU | Hyati pro | Yellow | NDD | - |
| 233 | SU | Hyati pro | Yellow | NDD | - |
| 234 | SU | Hyati pro | Yellow | NDD | - |
| 235 | SU | Hyati pro | Yellow | NDD | - |
| 236 | SU | Hyati pro | Yellow | NDD | - |
| 237 | SU | Hyati pro | Yellow | NDD | - |
| 238 | SU | Hyati pro | Yellow | NDD | - |
| 239 | SU | Hyati pro | Yellow | NDD | - |
| 240 | SU | Hyati pro | Yellow | NDD | - |
| 241 | SU | Hyati pro | Yellow | NDD | - |
| 242 | SU | Hyati pro | Yellow | NDD | - |
| 243 | SU | Hyati pro | Yellow | NDD | - |
| 244 | SU | Hyati pro | Yellow | NDD | - |
| 245 | SU | Hyati pro | Yellow | NDD | - |
| 246 | SU | Hyati pro | Yellow | NDD | - |
| 247 | SU | Ene Legend | Brown | NDD | - |
| 248 | SU | Ene Legend | Yellow | NDD | - |
| 249 | SU | Elux Legend | Brown | NDD | - |

| | | | | | |
|---|---|---|---|---|---|
| 250 | SU | Elux Legend | Brown | NDD | - |
| 251 | SU | Elux Legend | Brown | NDD | - |
| 252 | SU | Crystal bar | Yellow | NDD | - |
| 253 | SU | Crystal bar | - | NT | - |
| 254 | SU | Crystal prime | Yellow | NDD | - |
| 255 | SU | Randm Tornado | - | NT | - |
| 256 | SU | Randm Tornado | Brown | MDMB-4en-PINACA | 0.05 |
| 257 | SU | Elfbar | Brown | NDD | - |
| 258 | SU | Elfbar | Yellow | NDD | - |
| 259 | SU | Elfbar | Brown | NDD | - |
| 260 | SU | Elfbar | Brown | NDD | - |
| 261 | SU | Elfbar | Brown | NDD | - |
| 262 | SU | Crystal bar | Yellow | NDD | - |
| 263 | SU | Crystal bar | Yellow | NDD | - |
| 264 | SU | Crystal bar | Yellow | NDD | - |
| 265 | SU | - | - | NT | - |
| 266 | SU | - | Brown | NDD | - |
| 267 | SU | Crystal pro | Brown | NDD | - |
| 268 | SU | WGA | Yellow | NDD | - |
| 269 | SU | WGA | - | NT | - |
| 270 | SU | Hyati pro ultra | Yellow | NDD | - |
| 271 | SU | Lost mary | Green | NDD | - |
| 272 | SU | Lost mary | Yellow | NDD | - |

NDD, no drug detected; 'low', concentration below level of NMR measurement; IM, insufficient material; NT, not tested; SU, single use; RF, refillable; UB, unlabelled bottle; LB, labelled bottle.

**Table S3. Results summary for all samples in R3**

| R3S1 | Sample type | Detail | Liquid colour | Presumptive result |
|---|---|---|---|---|
| 1 | SU | Boutiq, French Toast | Yellow resin | THC |
| 2 | SU | CCELL | Yellow resin | THC |
| 3 | SU | Bubbglegum Runtz | Yellow resin | THC |
| 4 | RF | Argus | Yellow | SC |
| 5 | SU | Crystal bar | - | NDD |
| 6 | RF | Gotek | - | NDD |
| 7 | RF | Sonder | - | NDD |
| 8 | RF | Vaporesso | - | NDD |
| 9 | RF | Vaporesso | - | NDD |
| 10 | RF | Vaporesso | - | NDD |
| 11 | SU | Crystal prime | - | NDD |
| 12 | SU | Crystal prime | - | NDD |
| 13 | SU | Hyati pro, blue razz cherry | - | NDD |
| 14 | LB | Hyati, berry lemonade | - | NDD |
| 15 | LB | Hyati, strawberry watermelon | - | NDD |
| R3S2 | Sample type | Detail | Liquid colour | Presumptive result |
| 16 | RF | Vaporesso | Yellow | SC |
| 17 | RF | Vaporesso | Blue | SC |
| 18 | UB | - | Blue | SC |
| 19 | UB | - | Blue | SC |
| 20 | UB | - | Blue | SC |
| 21 | RF | Gotek | - | NDD |
| 22 | SU | Elfbar | - | NDD |
| 23 | RF | Sonder | - | NDD |
| 24 | RF | Vaporesso | - | NDD |
| 25 | RF | Vaporesso | - | NDD |
| 26 | LB | Hit Liquid | - | NDD |
| 27 | LB | Elux, berry lemonade | - | NDD |
| 28 | LB | Elux, gummy bear | - | NDD |
| 29 | SU | Hyati, Cherry cola | - | NDD |
| 30 | SU | Hyati, blue soure raspberry | - | NDD |
| 31 | SU | Hyati, triple mango | - | NDD |
| 32 | SU | Lux pro, Mr blue | - | NDD |
| 33 | SU | Hyati, strawberry raspberry ice | - | NDD |
| 34 | SU | Hyati | - | NDD |
| 35 | SU | Crystal prime | - | NDD |
| 36 | SU | Crystal prime | - | NDD |
| 37 | SU | Hyati ultra, blue razz gummy bear | - | NDD |
| 38 | SU | Hyati duo, rainbow sherbert | - | NDD |
| 39 | SU | Crystal, juicy peach | - | NDD |
| 40 | SU | Crystal pro plus | - | NDD |
| 41 | SU | Hyati, mad blue | - | NDD |
| R3S3 | Sample type | Detail | Liquid colour | Presumptive result |
| 42 | UB | - | Clear | SC |

| | | | | |
|---|---|---|---|---|
| 43 | UB | - | Clear | SC |
| 44 | UB | - | Pink | SC |
| 45 | UB | - | Yellow | SC |
| 46 | UB | - | Yellow | SC |
| 47 | UB | - | Yellow | SC |
| 48 | LB | Elux, gummy bear | - | NDD |
| R3S4 | Sample type | Detail | Liquid colour | Presumptive result |
| 49 | RF | Gotek | Green | SC |
| R3S5 | Sample type | Detail | Liquid colour | Presumptive result |
| 50 | RF | Backpackboyz | Yellow resin | THC |
| 51 | RF | CCELL | Yellow | THC |
| 52 | RF | Gotek | Green | SC |
| 53 | SU | Elfbar | - | NDD |
| 54 | RF | Vaporesso | - | NDD |
| 55 | LB | Gost red | - | NDD |
| 56 | LB | Elux, Gummey bear | - | NDD |
| 57 | LB | Bar juice, blueberry soure raspberry | - | NDD |
| 58 | LB | Pod salt nexus | - | NDD |
| 59 | RF | Vaporesso | - | NDD |
| 60 | RF | Vaporesso | - | NDD |
| 61 | RF | Gotek | - | NDD |
| 62 | RF | Gotek | - | NDD |
| 63 | RF | Powtop | - | NDD |
| 64 | RF | Vaporesso | - | NDD |
| 65 | RF | Vaporesso | - | NDD |
| 66 | RF | Gotek | - | NDD |
| 67 | RF | Gotek | - | NDD |
| 68 | RF | Gotek | - | NDD |
| 69 | RF | Gotek | - | NDD |
| 70 | SU | Randm Tornado | - | NDD |
| 71 | SU | Randm Tornado | - | NDD |
| 72 | SU | Randm Tornado | - | NDD |
| 73 | SU | Crystal bar | - | NDD |
| 74 | SU | Crystal bar | - | NDD |
| 75 | SU | Crystal bar | - | NDD |
| 76 | SU | Crystal bar | - | NDD |
| 77 | SU | Crystal prime | - | NDD |
| 78 | SU | Hyati pro, blue razz cherry | - | NDD |
| 79 | SU | Hyati pro, blue razz cherry | - | NDD |
| 80 | SU | Hyati pro ultra | - | NDD |
| 81 | SU | Hyati | - | NDD |
| 82 | SU | Hyati | - | NDD |
| 83 | SU | Hyati | - | NDD |
| 84 | SU | Elux | - | NDD |

NDD, no drug detected; SC, synthetic cannabinoid; THC, tetrahydrocannabinol; SU, single use; RF, refillable; UB, unlabelled bottle; LB, labelled bottle.

**Table S4. Data summary for R1, R2 and R3 for SC positives.**

| School | SC detected | Total submission | Total single use | Total liquid/refillable | Total alarm | Alarm single use | Alarm liquid/refillable | [Average] (mg / mL) | Alarm rate total | Alarm rate single use | Alarm rate liquid/refillable |
|---|---|---|---|---|---|---|---|---|---|---|---|
| R1S1 | Yes | 9 | 0 | 9 | 5 | 0 | 5 | 0.55 | 0.56 | 0.00 | 0.56 |
| R1S2 | Yes | 5 | 2 | 3 | 3 | 0 | 3 | 1.37 | 0.60 | 0.00 | 0.60 |
| R1S3 | No | 1 | 1 | 0 | 0 | 0 | 0 | 0.00 | 0.00 | 0.00 | 0.00 |
| R1S4 | Yes | 5 | 4 | 1 | 1 | 0 | 1 | 0.56 | 0.20 | 0.00 | 0.20 |
| R1S5 | No | 3 | 3 | 0 | 0 | 0 | 0 | 0.00 | 0.00 | 0.00 | 0.00 |
| R1S6 | Yes | 4 | 2 | 2 | 2 | 0 | 2 | 0.40 | 0.50 | 0.00 | 0.50 |
| R1S7 | Yes | 26 | 23 | 3 | 3 | 0 | 3 | 2.04 | 0.12 | 0.00 | 0.12 |
| R1S8 | Yes | 36 | 16 | 20 | 10 | 1 | 9 | 1.40 | 0.28 | 0.03 | 0.25 |
| R1S9 | Yes | 6 | 1 | 5 | 4 | 0 | 4 | 1.16 | 0.67 | 0.00 | 0.67 |
| R1S10 | No | 21 | 21 | 0 | 0 | 0 | 0 | 0.00 | 0.00 | 0.00 | 0.00 |
| R1S11 | Yes | 1 | 0 | 1 | 1 | 0 | 1 | 1.50 | 1.00 | 0.00 | 1.00 |
| R1S12 | Yes | 33 | 27 | 6 | 6 | 0 | 6 | 1.25 | 0.18 | 0.00 | 0.18 |
| R1S13 | No | 6 | 6 | 0 | 0 | 0 | 0 | 0.00 | 0.00 | 0.00 | 0.00 |
| | | **156** | **106** | **50** | **35** | **1** | **34** | **0.00** | **0.224** | **0.01** | **0.22** |
| R2S1 | Yes | 31 | 24 | 7 | 12 | 6 | 6 | 0.88 | 0.39 | 0.19 | 0.19 |
| R2S2 | Yes | 32 | 20 | 12 | 3 | 0 | 3 | 0.34 | 0.09 | 0.00 | 0.09 |
| R2S3 | Yes | 99 | 71 | 28 | 13 | 0 | 13 | 0.76 | 0.13 | 0.00 | 0.13 |
| R2S4 | Yes | 3 | 0 | 3 | 2 | 0 | 2 | 0 | 0.67 | 0.00 | 0.67 |
| R2S5 | Yes | 25 | 20 | 5 | 1 | 0 | 1 | 0 | 0.04 | 0.00 | 0.04 |
| R2S6 | Yes | 6 | 3 | 3 | 3 | 0 | 3 | 1.86 | 0.50 | 0.00 | 0.50 |
| R2S7 | No | 7 | 4 | 3 | 0 | 0 | 0 | 0 | 0.00 | 0.00 | 0.00 |
| R2S8 | No | 13 | 12 | 1 | 0 | 0 | 0 | 0 | 0.00 | 0.00 | 0.00 |
| R2S9 | Yes | 54 | 42 | 12 | 6 | 1 | 5 | 0.47 | 0.11 | 0.02 | 0.09 |
| | | **270** | **196** | **74** | **40** | **7** | **33** | **0** | **0.148** | **0.03** | **0.12** |

| School | SC detected | Total submission | Total single use | Total liquid/refillable | Total alarm | Alarm single use | Alarm liquid/refillable | [Average] (mg / mL) | Alarm rate total | Alarm rate single use | Alarm rate liquid/refillable |
|---|---|---|---|---|---|---|---|---|---|---|---|
| R3S1 | Yes | 15 | 7 | 8 | 1 | 0 | 1 | NA | 0.07 | 0.00 | 0.07 |
| R3S2 | Yes | 26 | 14 | 12 | 5 | 0 | 5 | NA | 0.19 | 0.00 | 0.19 |
| R3S3 | Yes | 7 | 0 | 7 | 6 | 0 | 6 | NA | 0.86 | 0.00 | 0.86 |
| R3S4 | Yes | 1 | 0 | 1 | 1 | 0 | 1 | NA | 1.00 | 0.00 | 1.00 |
| R3S5 | Yes | 35 | 16 | 19 | 1 | 0 | 1 | NA | 0.03 | 0.00 | 0.03 |
| | | 84 | 37 | 47 | 14 | 0 | 14 | NA | 0.167 | 0.00 | 0.167 |
| | **0.75** | **510** | **339** | **171** | **89** | **8** | **81** | **1.03** | **0.1745** | **0.0157** | **0.159** |

**Table S5. Results summary for all samples in R4.**

| R4S7 | Sample type | Detail | Liquid colour | GC-MS result |
|---|---|---|---|---|
| 1 | SU | crystal prime | - | NDD |
| 2 | SU | Jeeter | - | THC |
| 3 | RF | ox | - | 4F-MDMB-BINACA |
| 4 | SU | novo | - | 4F-MDMB-BINACA |
| 5 | SU | non refillable firerose | - | NDD |
| 6 | RF | red oxva | - | 4F-MDMB-BINACA |
| 7 | RF | blue oxva | - | 4F-MDMB-BINACA |
| 8 | SU | happy vibes | - | NDD |
| 9 | SU | green crystal prime | - | NDD |
| 10 | LB | elux vape refill blueberry | - | NDD |
| 11 | LB | jucce candy black jack | - | NDD |
| 12 | UB | unknown -in small vial no ID | Clear | MDMB-4en-PINACA |
| 13 | LB | pukka juice lime lemonade | - | NDD |
| 14 | LB | soda king lemon lime | - | NDD |
| 15 | LB | crystal clear blueberry sour raspberry | - | NDD |
| 16 | RF | - | - | MDMB-4en-PINACA |
| 17 | RF | OXVA gold/green with refill top | - | NDD |
| 18 | LB | elux vape refill strawberry ice cream | - | NDD |
| 19 | LB | elux vape refill kiwi passion fruit | - | NDD |
| 20 | LB | elux vape refill cherry cola | - | NDD |
| 21 | RF | unknown | - | NDD |
| 22 | RF | black and gold | - | MDMB-4en-PINACA |
| 23 | SU | pale green KLAK | - | NDD |
| 24 | RF | lime green and green OXVA | - | NDD |
| 25 | LB | elux tiger blood | - | NDD |
| 26 | RF | blue and black OXVA | - | NDD |
| 27 | LB | elflio - cherry | - | NDD |
| 28 | RF | silver oxva | - | MDMB-4en-PINACA |
| 29 | SU | - | - | NDD |
| 30 | RF | - | - | MDMB-4en-PINACA |
| 31 | RF | - | - | NDD |
| 32 | RF | - | - | NDD |
| 33 | RF | - | - | MDMB-4en-PINACA |
| 34 | UB | - | Pink | MDMB-4en-PINACA |
| 35 | RF | - | - | MDMB-4en-PINACA |
| 36 | RF | - | - | NDD |
| 37 | RF | - | - | MDMB-4en-PINACA |
| 38 | RF | - | - | NDD |
| 39 | RF | - | - | MDMB-4en-PINACA |
| 40 | RF | - | - | MDMB-4en-PINACA |
| 41 | LB | mango | - | NDD |
| 42 | LB | watermelon | - | NDD |
| 43 | LB | blueberry | - | NDD |
| 44 | UB | - | green | MDMB-4en-PINACA |

| | | | | |
|---|---|---|---|---|
| 45 | SU | yellow with white top | - | THC |
| 46 | SU | crystal fire blue and yellow | - | NDD |
| 47 | LB | lemon and lime nearly full | - | NDD |
| 48 | LB | lemon and lime nearly empty | - | NDD |
| 49 | SU | lost mary red/yellow | - | NDD |
| 50 | SU | Hayati pro max | - | NDD |

NDD, no drug detected; THC, tetrahydrocannabinol; SU, single use; RF, refillable; UB, unlabelled bottle; LB, labelled bottle.

**Table S6. LC-MS parent molecule and fragment ion mass to charge (mz) ratios used to confirm compounds found in the e-cigarettes/liquids.**

| Compound | Parent | F1 | F2 | F3 | F4 | F5 | F6 |
|---|---|---|---|---|---|---|---|
| MDMB-4en-PINACA | 358.2125 | 213.1022 | 298.1914 | 145.0396 | 86.0964 | 171.089 | 163.0502 |
| ADB-BUTINACA | 331.2128 | 201.1022 | 286.1914 | 314.1863 | 163.0502 | 145.0396 | 219.1128 |
| 4F-MDMB-BINACA | 364.2031 | 219.0928 | 304.1819 | 145.0396 | 237.1034 | 237.1022 | 236.1193 |
| MDMB-PINACA | 360.2281 | 215.1179 | 145.0396 | 300.207 | | | |
| ADB-4en-PINACA | 343.2128 | 213.1022 | 298.1914 | 326.1863 | 365.1948 | | |
| MDMB-INACA | 290.1499 | 145.0396 | 230.1288 | 86.0964 | | | |
| Δ9-THC | 315.2131 | 193.1223 | 123.0441 | 259.1693 | 235.1693 | 233.1536 | 221.1536 |

## R1S1.2

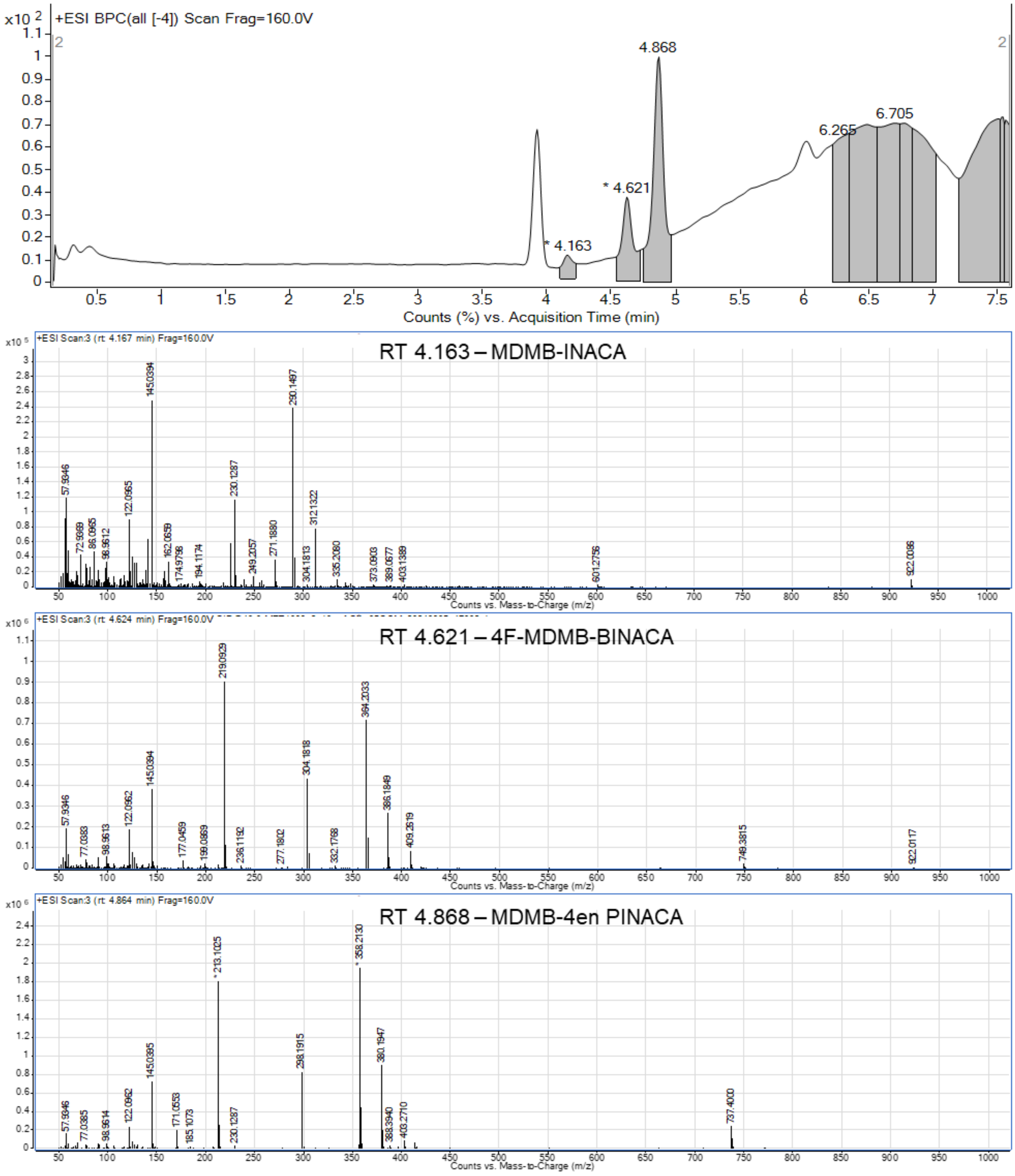


**Figure S1.** Example LC-MS chromatogram and spectra for sample R1S1.2 with MDMB-4en-PINACA, 4F-MDMB-BINACA and MDMB-INACA SCs identified. Molecular weights of compounds: MDMB-INACA – 289.330 g $mol^{-1}$, 4F-MDMB-BINACA – 363.433 g $mol^{-1}$, MDMB-4en-PINACA - 357.454 g $mol^{-1}$.

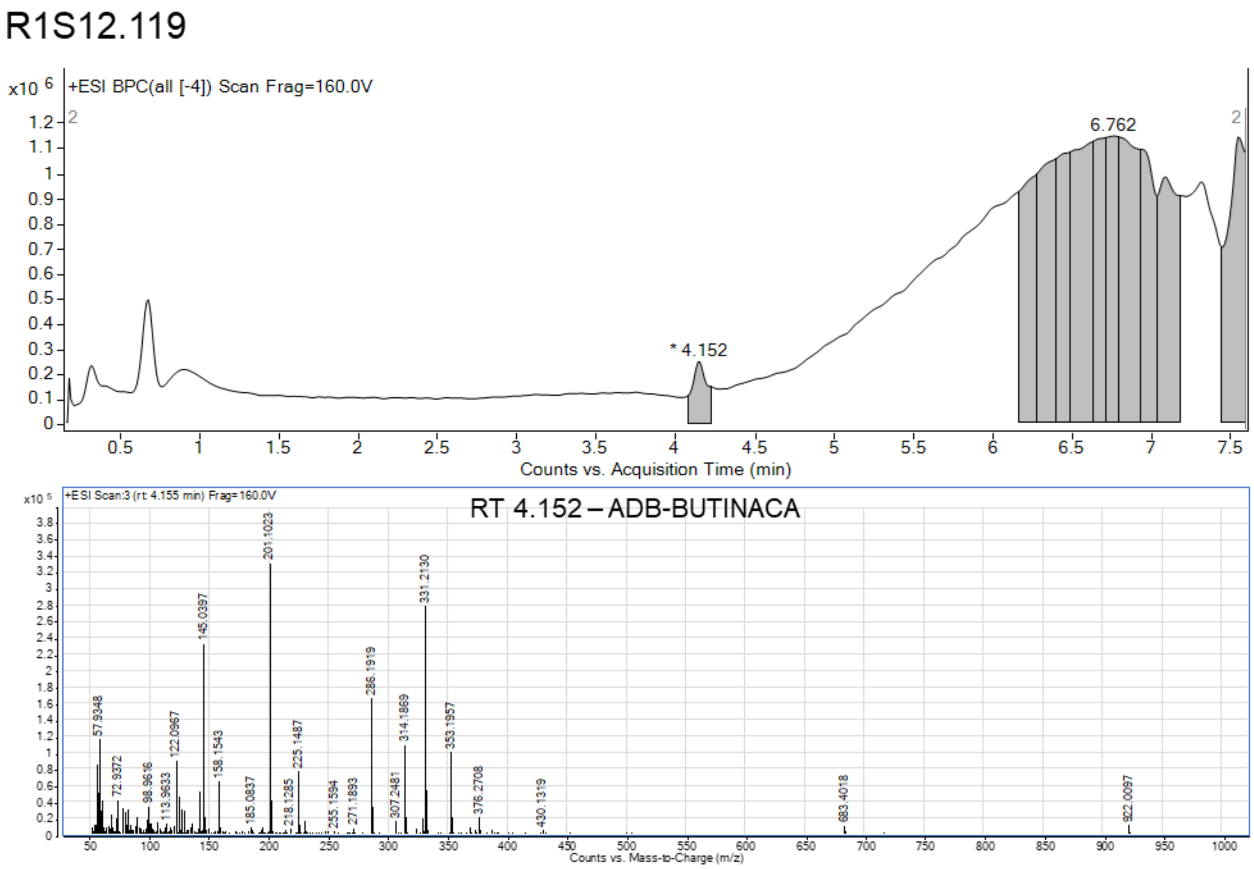


**Figure S2.** Example LC-MS chromatogram and spectra for sample R1S12.119 with ADB-BUTINACA identified. Molecular weight of ADB-BUTINACA is 330.432 g $mol^{-1}$.

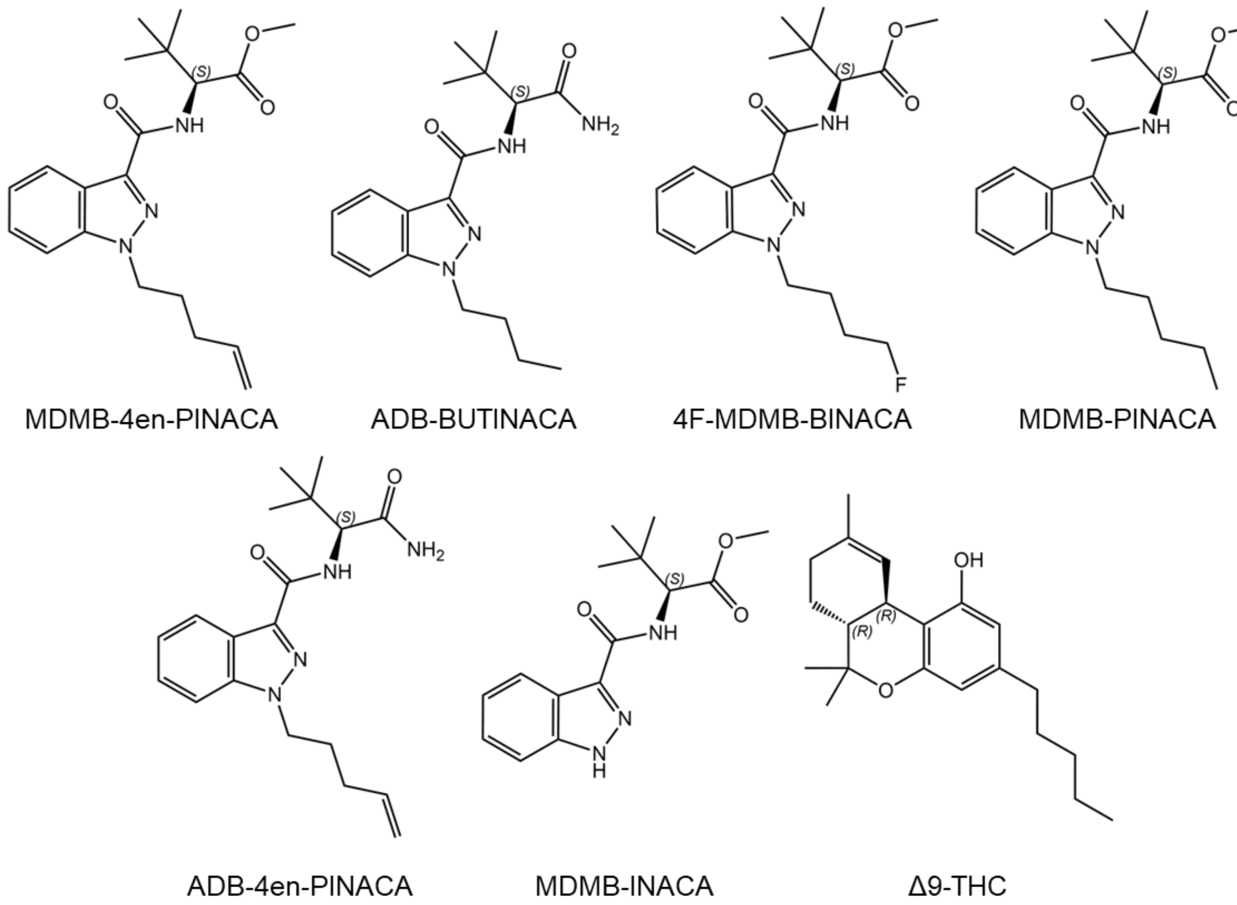


**Figure S3.** Structures of illicit drugs identified.

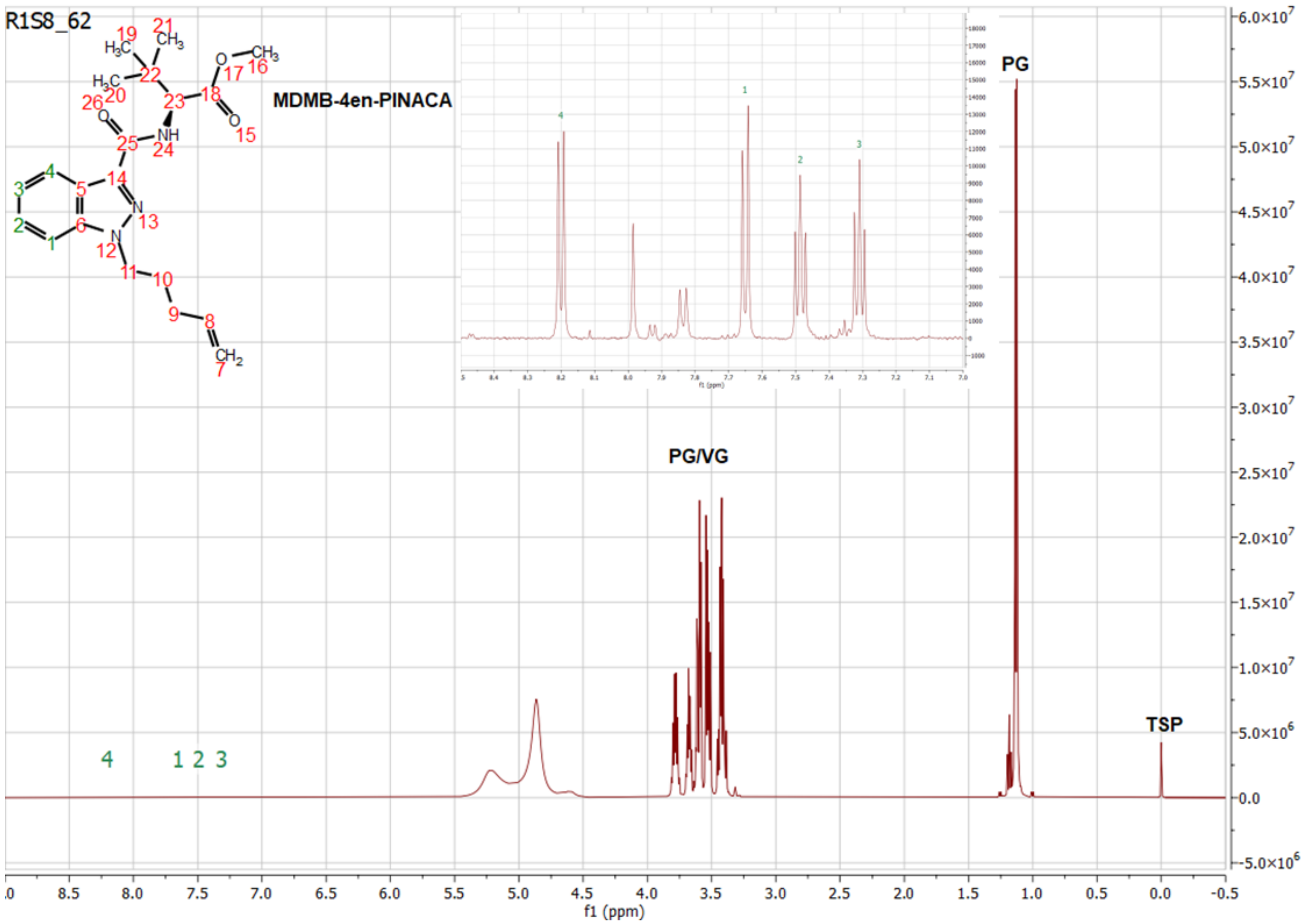


**Figure S4.** Example 1H NMR spectra used for qNMR on sample R1S8.62 containing MDMB-4en-PINACA. Main spectra shows the sample is dominated by polyethylene glycol (PG) and vegetable glycerin (VG) that form the basis of e-cigarette liquid. Insert is of the 8.5 – 7.0 ppm region which shows the aromatic indazole peaks for hydrogens 1-4 used for the quantification. The internal standard 3-(trimethylsilyl)propionic-2,2,3,3-d4 acid sodium salt (TSP) is shown at 0.0 ppm (3 mg added). The qNMR method used a 20 s delay, 128 scans and processing techniques (as described in materials and methods) such that ratio of the integration of the sample to TSP peaks can be used to calculate the concentration of the sample, 3.53 mg/mL in this case.

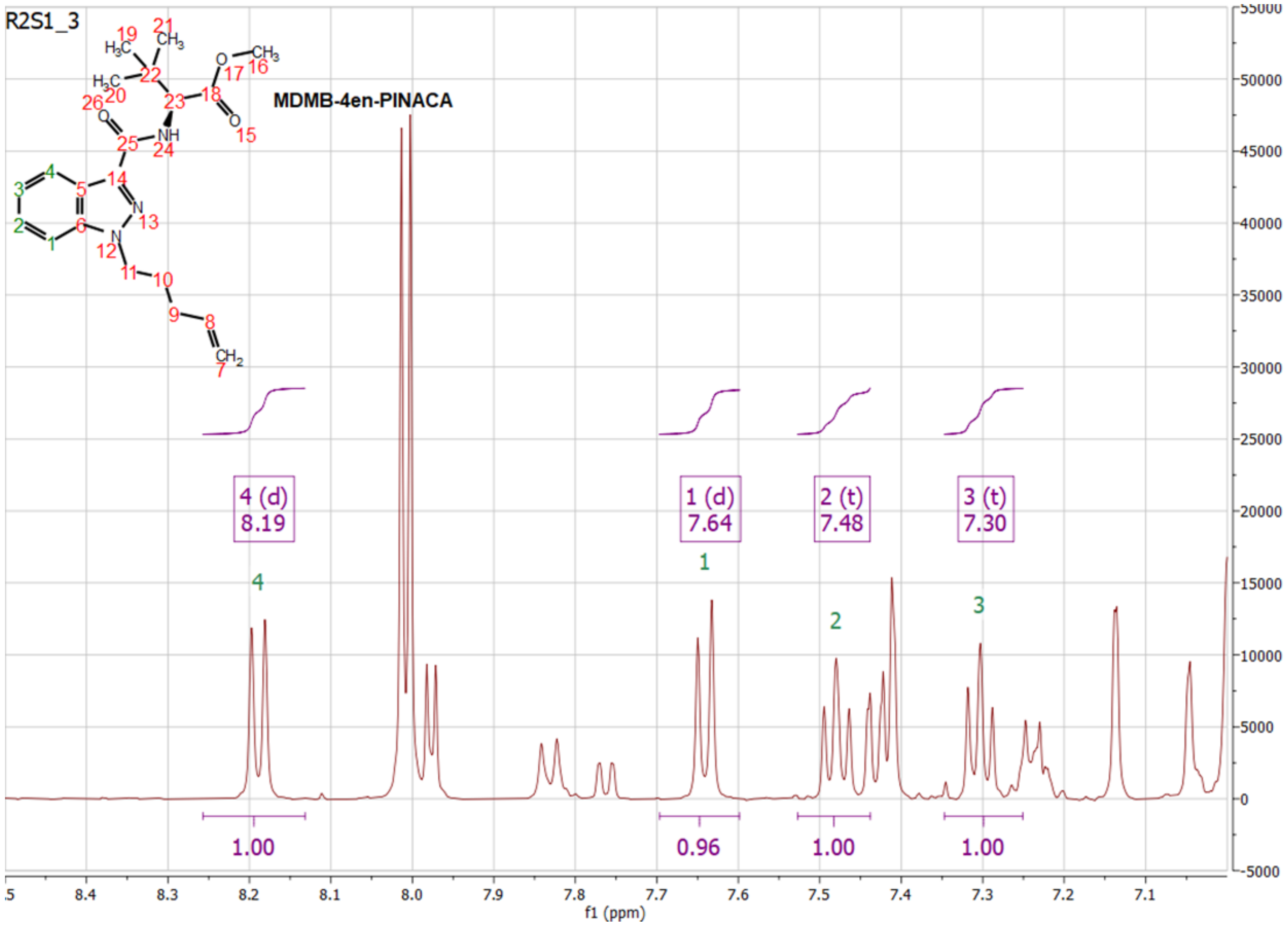


**Figure S5.** Example 1H NMR spectra used for qNMR on sample R2S1.3 containing MDMB-4en-PINACA. The aromatic indazole 8.5 – 7.0 ppm region is shown with the multiplets and integration of hydrogens 1-4 used for the quantification.

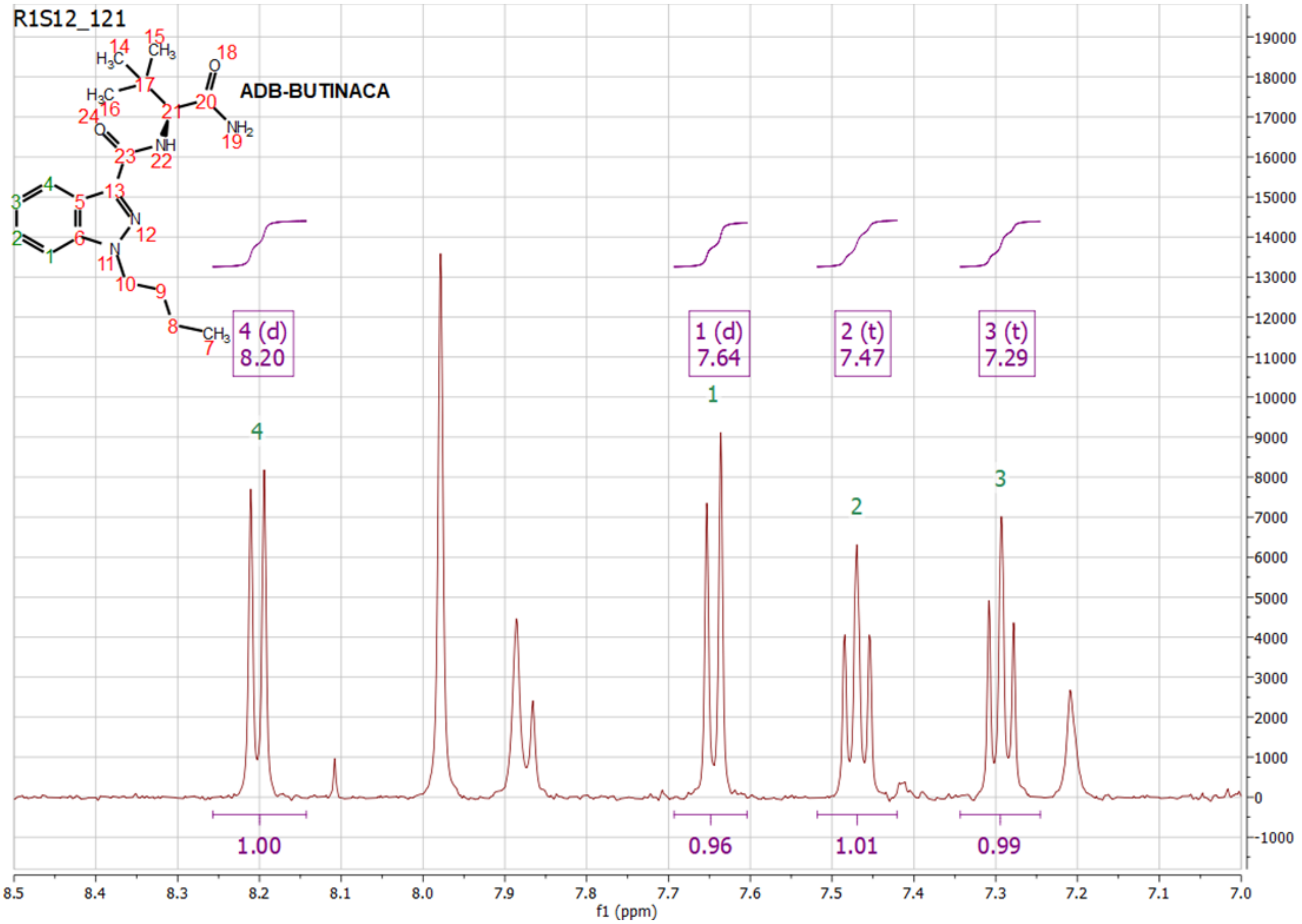


**Figure S6.** Example 1H NMR spectra used for qNMR on sample R1S12.121 containing ADB-BUTINACA. The aromatic indazole 8.5 – 7.0 ppm region is shown with the multiplets and integration of hydrogens 1-4 used for the quantification.

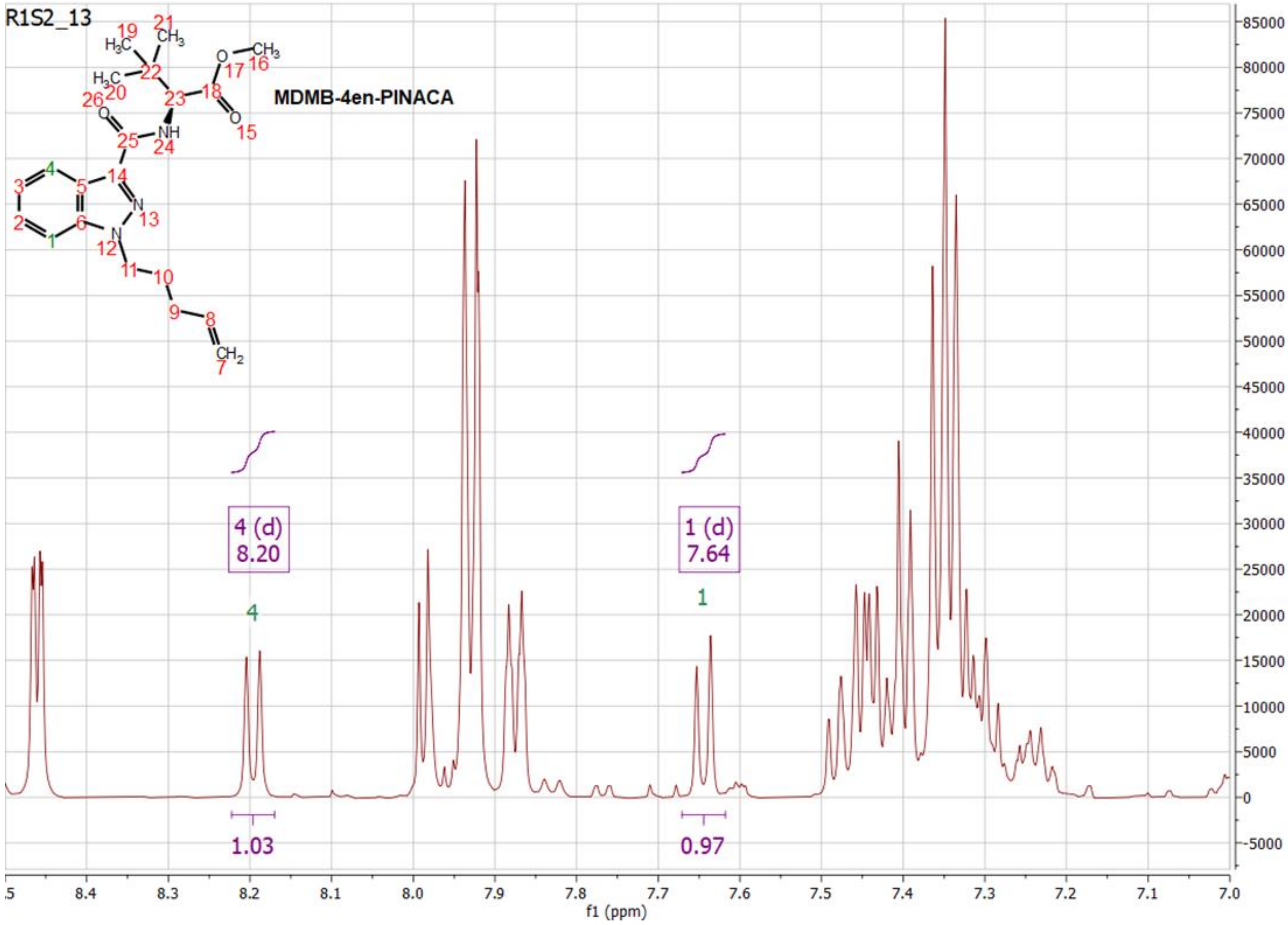


**Figure S7.** Example 1H NMR spectra used for qNMR on sample R1S2.13 containing MDMB-4en-PINACA. The aromatic indazole 8.5 – 7.0 ppm region is shown highlighting that only the multiplets and integration of hydrogens 1 and 4 are able to be used for quantification due to additives in the e-cigarette liquid masking the other hydrogens.

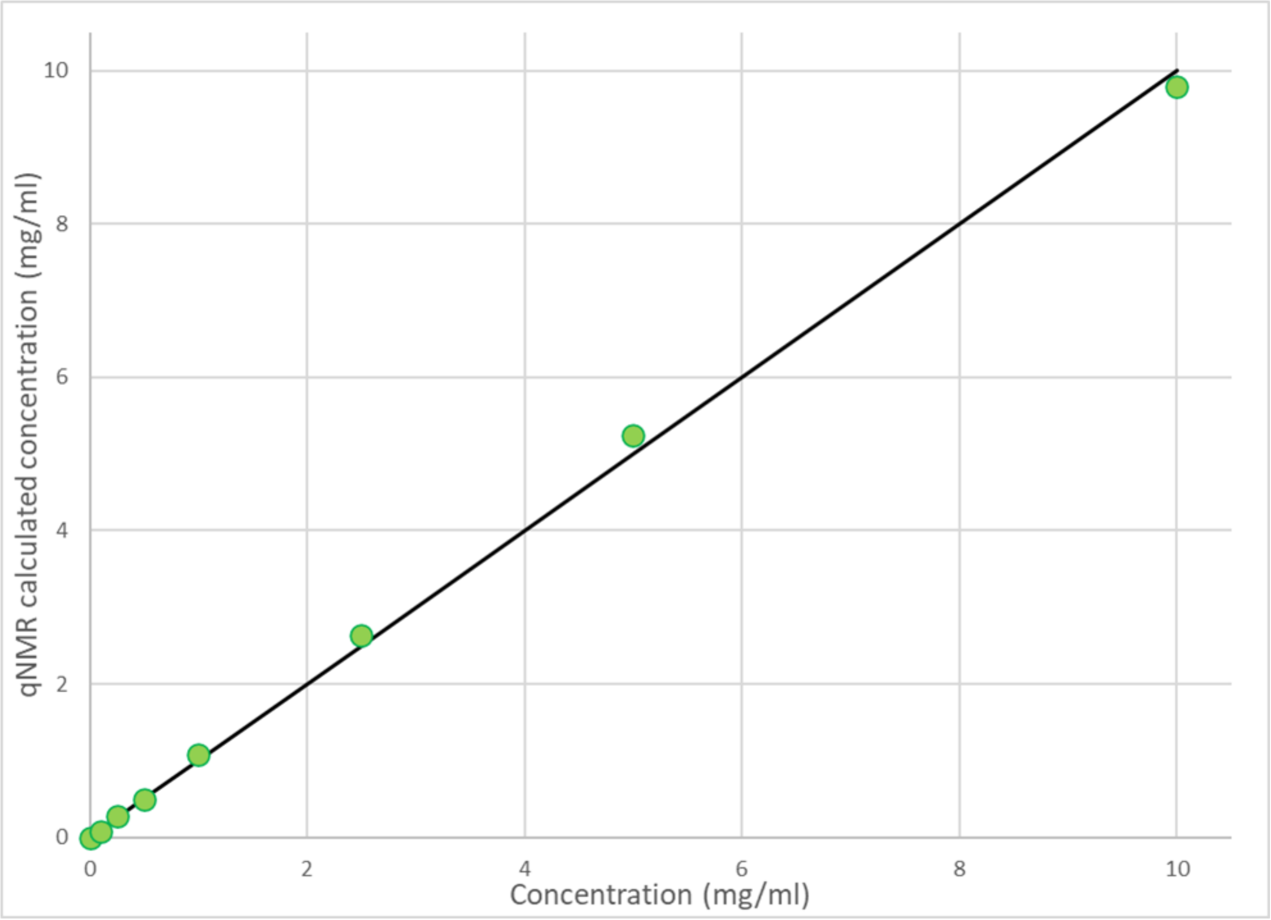


**Figure S8.** Plot showing the qNMR calculated values of a standard concentration range of MDMB-4en-PINACA in e-cigarette liquid. This shows good agreement with the true values (black line) highlighting the suitability of the qNMR technique for quantifying the e-cigarettes and liquids.

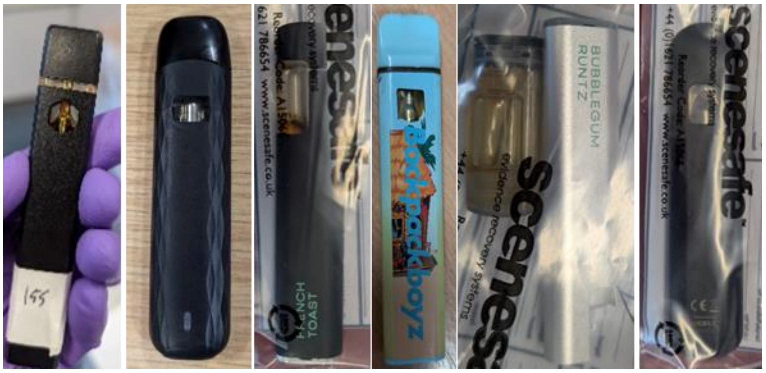


**Figure S9.** THC e-cigarettes from R3.

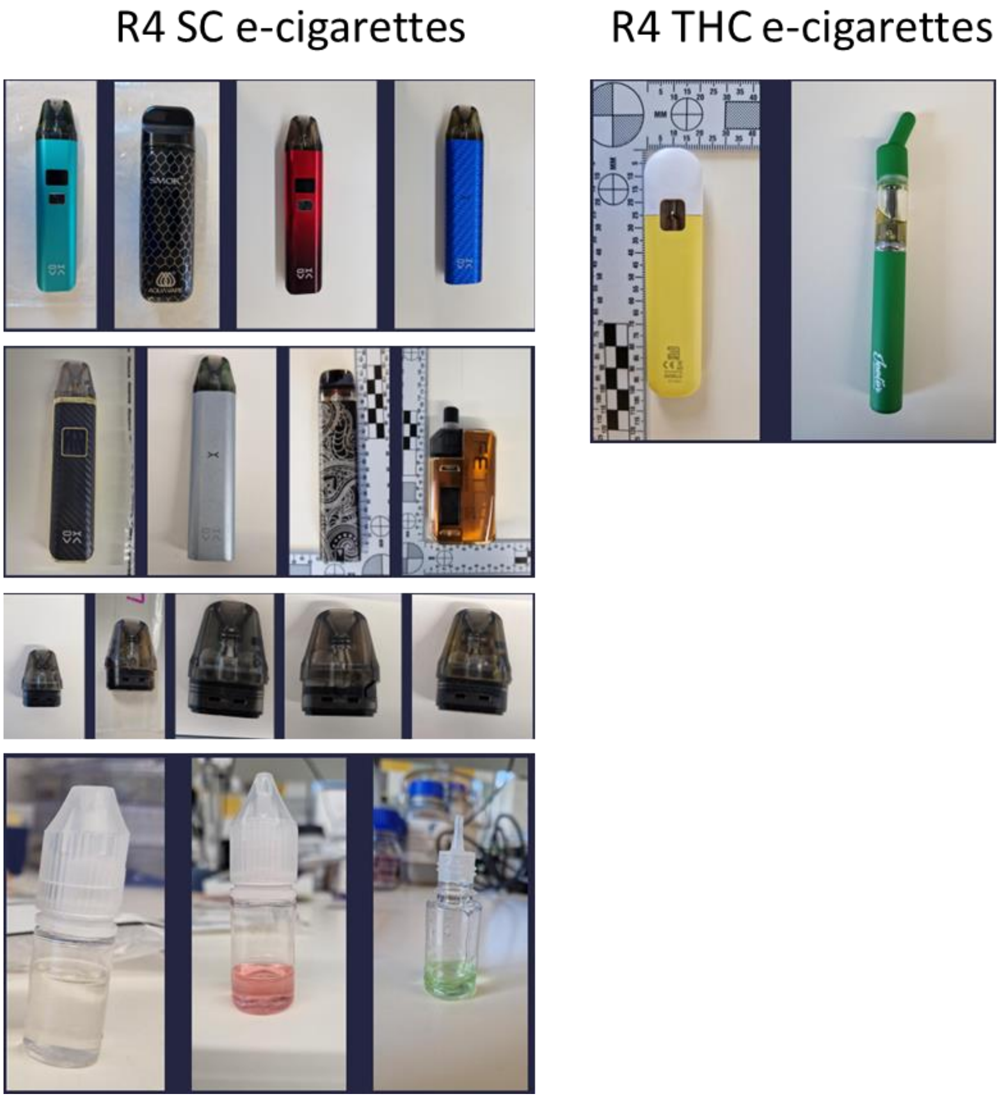


**Figure S10.** Positive samples (SC and THC) from R4.
